# Projected effects of removing pharmacists from pharmacies on access, prices and medication safety in Chile

**DOI:** 10.64898/2026.11.23.26368576

**Authors:** Carlos Balmaceda, Juan Carlos Ríos, Daniela Lahoz, Manuel Espinoza

## Abstract

**Background:** Chilean law requires a pharmacist to direct every pharmacy throughout opening hours. Bill 18,719-11 would allow any pharmacy to replace the pharmacist with another health professional or a technician, arguing that cheaper direction would open pharmacies where there are none and lower prices. We assessed that argument and the safety consequences.

**Methods:** We built a societal-perspective simulation model comparing the bill with current regulation over 2027–2036. Pharmacist hours replaced determine the share of dispensings without a pharmacist, which drives additional adverse drug reactions and therapeutic-error poisonings, estimated from national hospital discharges and published data of the national poison centre. New access in municipalities without a pharmacy was valued. The increase in errors without a pharmacist and the share of harm originating in the affected pharmacies are unmeasured and were treated as scenarios. Labour-cost savings and the net health and access value were reported separately, discounted at 3%.

**Results:** In the reference scenario, 27% of dispensings would occur without a pharmacist by 2036. Over ten years the bill would add about 2,500 admissions for adverse drug reactions, 670 poisonings from therapeutic errors (28 of them admitted) and 68 deaths (95% interval 22–166). About 17,700 people would gain a pharmacy, worth CLP 12 billion (US$13 million), against CLP 150 billion (US$165 million) in health lost and care. The net health and access value was negative in 99.9% of simulations, and with any increase in errors above 3.2%. Pharmacies would save CLP 270 billion (US$296 million) in pay, lowering medicine spending by at most 0.6–1.9% if fully passed on. A favourable economic balance would require nearly three-quarters of displaced pharmacists to be re-employed immediately at the same pay.

**Conclusions:** The access and price gains are small and local, whereas the health risks would be national. They do not justify allowing every pharmacy to replace its pharmacist; access in isolated localities calls for targeted measures.

## Introduction

Many countries require a pharmacist to direct community pharmacies or to be present when medicines are dispensed. The rationale is an information asymmetry: patients cannot judge whether a dose is correct or whether medicines interact. A professional requirement corrects that asymmetry but makes the service more expensive.^1^

In Chile, every pharmacy must have a pharmacist as technical director, present during all opening hours. In municipalities with no pharmacy, a drugstore (almacén farmacéutico) run by a technician or a pharmacy assistant may sell a restricted list that excludes psychotropics and narcotics.^2^ Bill 18,719-11, introduced in September 2026, would allow any pharmacy in the country to be directed by another health professional or by a higher-level health technician trained in pharmacology and dispensing.^3^ The director must be present and may sell every medicine. Its sponsors argue that the pharmacist’s salary makes small outlets unviable in some 50 municipalities and that cheaper direction would increase competition and lower prices, but present no data on either effect. An earlier bill proposed an exception limited to isolated localities.^4^

The argument is a chain: cheaper direction allows more outlets, more outlets increase competition, competition lowers prices and lower prices improve access. Evidence on each link is partial. European deregulation added pharmacies mostly in urban areas that already had them.^5^ In Portugal, supermarket entry lowered over-the-counter prices by 4–6%.^6^ In Chile, municipal pharmacies raised nearby private prices but lowered total spending.^7^ On safety, wider sale of paracetamol in Sweden was associated with more poisonings,^8^ and the Chilean poison centre recorded 15,077 medication errors in 2019–2024, mostly at home, with tramadol the medicine most often involved.^9^ No study has measured what happens when a pharmacist is replaced by a non-pharmacist director in the same pharmacy.

Chile has about 4,600 pharmacies; in 2015–2018 three chains held 42% of outlets and 80% of sales.^10^ Some 55 municipalities, with more than 300,000 residents, have no pharmacy of any kind.^11^ Much of the treatment for hypertension, diabetes and dyslipidaemia is dispensed by the public network rather than by private pharmacies.^12^ Similar debates arise across Latin America, where several countries require a pharmacist to direct each pharmacy while others allow other directors in some outlets. We asked how much access and price reduction the bill can deliver, what health harm replacing pharmacists could cause, and under which conditions the bill would be better or worse for society than current regulation.

## Methods

### Study design and comparators

We compared current regulation with the proposal, under which any pharmacy may replace its pharmacist with a technician, paid the current market wage (a 20% premium for the director’s legal responsibility is a variant). Current regulation is held constant, so results are differences from it. Unmeasured quantities, such as how many pharmacists would be replaced and how much errors would increase without them, were treated as scenarios (Table 1). The evaluation is ex ante, over 2027–2036, in 2026 Chilean pesos (CLP; US$1 = CLP 912). Following national guidance, costs and outcomes were discounted at 3% a year,^13^ with 0% and the official social rate of 5.5% as sensitivity analyses, and a statistical life was valued at CLP 2,384 million.^14^ Unlike the guidance, we took a societal perspective, because the bill’s effects fall on pharmacy owners, workers and patients, not on the health budget alone. With *t* = 1 for 2027, present values are PV(*X*) = ∑_*t*_ *X*_*t*_/1.03 ^*t*−1^. Equations, parameters and sources are given in Supplementary Notes 1–3 and Supplementary Tables 2–10, and a worked example with base values in Supplementary Table 1.

**Table 1.** Scenarios for quantities that have not been measured.

| **Quantity** | **Meaning** | **Values evaluated** | **Basis** |
|---|---|---|---|
| Long-run share of pharmacist posts replaced | Chains (independents without an owner-pharmacist) | 25% (15%); 45% (30%), reference; 70% (50%) | Future behaviour of pharmacies; no evidence |
| Replacement rule | Which hours are replaced first | Weekend first, reference; proportional across all hours | Authors’ judgement; the bill sets no limit |
| Increase in errors reaching the patient without a pharmacist (Δ) | Relative increase in errors interceptable at dispensing (ADRs and therapeutic errors) | 0%; 20%; 39%, reference; 67% | Odds ratio of 0.60 chosen between two studies of antibiotic sales |
| Share of drug-related harm passing through the affected pharmacies (*s*<sub>*F*</sub>) | Share of ADR and therapeutic-error admissions originating in medicines they dispense | 25%; 50%, reference; 75%; 100% (equal weight) | No Chilean estimate |
| Therapeutic errors interceptable at the pharmacy | Share of therapeutic-error poisonings a pharmacy could prevent | 3.4% (strict); 24.8% (excluding self-medication); 44.3% (broad), reference | National poison centre, annual report 2024 |
| Case fatality of additional ADR admissions | Deaths per admission | 0.5%; 1%; 2.1%; 2.7%, reference; 3%; 5% | Chilean discharges 2019 and 2023, weighted by age |
| Productive recovery of the replaced pharmacist (*φ*) | Value produced in the new job, as a fraction of previous labour cost | 0% to 100%, no preferred value; 80% as comparison anchor | No Chilean data |
| Opportunity cost of the technician (*θ*) | Value of the technician’s previous work, as a fraction of labour cost | 100%, reference; 75%; 50% | Authors’ judgement |

*ADR, adverse drug reaction. Reference values are working values used to order the presentation; they are not the most likely values.*

**Table 2.** Results of the proposal relative to current regulation, reference scenario, 2027–2036.

| Outcome | Mean (95% interval) |
|---|---|
| **Exposure in 2036** |  |
| Full-time pharmacist posts replaced | 2,330 (1,650 to 3,180) |
| Opening hours without a pharmacist | 39% (36 to 41) |
| Dispensings without a pharmacist | 27% (23 to 31) |
| **Health outcomes, 2027–2036 (not discounted)** |  |
| Admissions for adverse drug reactions | 2,470 (838 to 5,993) |
| Emergency visits for adverse drug reactions, not admitted | 6,465 (2,060 to 16,128) |
| Survivors of adverse drug reactions with sequelae | 226 (62 to 588) |
| Poisonings from therapeutic errors | 666 (461 to 823) |
| Of which admitted | 28 (16 to 41) |
| Of which managed without admission | 638 (442 to 788) |
| Of which in children under 12 years | 304 |
| Deaths from adverse drug reactions | 67 (22 to 166) |
| Deaths from poisoning | 0.30 (0.11 to 0.63) |
| DALYs | 1,222 (400 to 3,035) |
| Of which years of life lost | 1,140 (369 to 2,830) |
| Of which years lived with disability | 82 (21 to 230) |
| Antibiotic courses sold without prescription (exploratory, not valued) | 219,000 (88,300 to 420,400) |
| **Access, 2027–2036** |  |
| People gaining a pharmacy in their municipality (from 2029) | 17,700 (2,800 to 45,400) |
| **Present value, 2027–2036 (CLP billion)** |  |
| Value of access | +12 (+2 to +38) |
| Health lost | −141 (−346 to −46) |
| Of which deaths valued at the VSL | −137 (−337 to −45) |
| Of which years lived with disability valued at the VSLY | −4 (−11 to −1) |
| Additional care costs, including carers’ time | −9 (−23 to −3) |
| = Net health and access value (access, health lost and care costs) | −138 (−357 to −34) |
| Labour-cost savings of pharmacies (not added) | +270 (+137 to +443) |

*Means (95% intervals) of 10,000 simulations. Medium replacement, weekend first, 39% increase in errors, half of the drug-related harm passing through the affected pharmacies, broad criterion for interceptable therapeutic errors. Children under 12 years apply the share reported by the poison centre for 2024 (45.6%). Monetary values are present values in 2026 Chilean pesos, discounted at 3%; CLP 1 billion = US\$1.1 million. DALY, disability-adjusted life year; VSL, value of a statistical life; VSLY, value of a statistical life-year.*

### Ethics

The study used only publicly available data: de-identified hospital discharge records published as open data by the Chilean Ministry of Health, aggregated statistics published by the national poison centre, and published literature. No individual was contacted and no data that are not publicly available were used, so ethical review was not required.

### Separating private savings from social value

Lower pay for the director saves the owner money but is a transfer, not a social gain. We therefore report three accounts that are not added together:

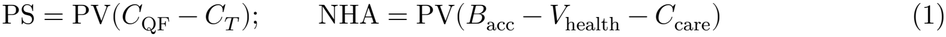

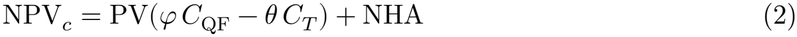

The labour-cost savings of pharmacies, PS, are the labour cost of the pharmacists released, *C*_QF_, minus that of the technicians hired, *C*_*T*_ ; they measure the owners’ incentive to replace and become profit unless passed on to prices. The net health and access value, NHA, adds the value of new access, *B*_acc_, and subtracts the value of health lost, *V*_health_, and the cost of additional care, *C*_care_. The complementary social net present value, NPV_*c*_, adds the social value of labour. A replaced pharmacist may produce elsewhere a fraction *φ* of the value of their previous work (productive recovery), and the technician leaves behind work worth a fraction *θ* of their labour cost (opportunity cost). Neither has been measured in Chile. We used *θ* = 1 as reference and 0.5 as sensitivity, and examined *φ* over its full range, fixing it at 0.8 only as an anchor when comparing other assumptions.

### Dispensing without a pharmacist

A pharmacy needs enough full-time pharmacist posts to cover its opening hours with one pharmacist present at a time. A chain outlet open 75 hours a week needs 75/40 = 1.9 posts, plus 6% cover for absences. A scenario sets the long-run share of posts replaced: 25%, 45% (reference) or 70% in chains, and 15%, 30% or 50% in independents not directed by their owner. We assumed, from the authors’ knowledge of the sector, that 82% of independents (range 75–90%) are directed by an owner-pharmacist, who would not be replaced. Half the long-run share is reached in the first year and half of the remainder each year after. Replacement is defined by who is present, not by pay.

With one pharmacist per hour, replacing 45% of a chain’s posts leaves 45% of its hours without one. The weekend is 27% of opening hours, and a weekend hour dispenses about 30% of a weekday hour (authors’ judgement, range 20–40%), so the weekend carries only 10% of dispensings. In the reference, weekend hours are replaced first, so a chain replacing 45% of its hours loses its pharmacist for the whole weekend (10% of dispensings) and a quarter of weekday hours (22%): 32% of its dispensings. Across existing pharmacies, the national share of dispensings made without a pharmacist, *E*_*F*_ , is

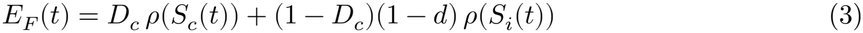

where *S*_*c*_ and *S*_*i*_ are the shares of hours replaced in chains and independents, *ρ*(*S*) the corresponding share of dispensings (equal to *S* under a proportional rule), *D*_*c*_ = 0.85 the chains’ share of dispensings and *d* = 0.82 the share of independents directed by their owner-pharmacist. In 2036, *E*_*F*_ ≈ 0.85 × 0.32 + 0.15 × 0.18 × 0.14 ≈ 0.277. New outlets in municipalities without a pharmacy add a small exposure, *D*_*N*_ (*t*), equal to their share of national dispensings.

### Adverse drug reactions

Public data do not identify drug-related admissions, so the baseline for community-acquired ADRs, *H*_0_, used age-specific rates from a systematic review: medians of 4.1% in children, 6.3% in adults and 10.7% in older adults.^15^ These were applied to 2019 non-surgical discharges from the national hospital discharge database of the Ministry of Health,^16^ the most recent year whose open data identify surgery, projected to 2026 with national population projections by age. The result was 67,695 admissions a year (95% interval 36,471–145,642), growing 1.7% a year with population ageing (*g*). Additional ADR admissions are

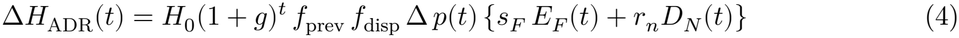

Here *f*_prev_ = 0.52 is the preventable fraction^17^ and *f*_disp_ = 0.15 the share of what is preventable that can be intercepted at dispensing, an assumption informed by a systematic review.^18^ *p*(*t*) phases the effect in from 50% in the first year to 100% by the fifth, and *r*_*n*_ = 1 counts dispensings by new outlets as made without a pharmacist.

Two factors in equation (4) are scenarios. The first, Δ, is the relative increase in interceptable errors reaching the patient when no pharmacist is present. The closest evidence concerns non-prescription antibiotic sales, which are lower when a pharmacist is present: odds ratios of 0.66 in China^19^ and 0.54 in Sri Lanka.^20^ We took 0.60, a value between the two, not a pooled estimate. With an assumed probability of 0.30 that a pharmacy sells antibiotics without prescription when a pharmacist is present, it converts to a relative increase of 1/(0.30 + 0.60 × 0.70) − 1 = 0.39. We evaluated Δ = 0 (no increase), 0.20 (low), 0.39 (reference) and 0.67 (high; the odds ratio read as a risk ratio). The second, *s*_*F*_ , is the share of drug-related admissions originating in medicines dispensed by the affected pharmacies rather than by primary care or hospitals; with no Chilean estimate, we evaluated 25%, 50% (reference), 75% and 100% with equal weight.

Each ADR admission generates 2.66 emergency visits that are not admitted,^21^ and 10% of survivors have sequelae. In Chilean discharges coded as adverse drug events in 2019 and 2023, in-hospital case fatality was 0% under 20 years, 1.2% at 20–59 years and 3.7% at 60 years or older. Weighted by the age mix of *H*_0_, 65% of which is aged 60 years or older, it is 2.7%, close to the 2.3% of a large British study;^22^ each death loses 16.9 years of life (Supplementary Note 6).

### Poisonings from therapeutic errors

Poisonings follow the same structure. Their baseline is the 670 annual admissions for accidental medicine poisoning (codes X40–X44), 20.8% of them therapeutic errors, the share among unintentional exposures reported to the national poison centre (CITUC) in its 2024 annual report.^23^ Of the therapeutic errors in that report, 44.3% were of types a pharmacy could intercept (Supplementary Table 23); this share is applied to admissions and to the 3,144 errors a year managed without admission, a lower bound because not every case reaches the centre. Since 2014, non-prescription medicines may be sold from open shelves without the pharmacist’s involvement,^24^ so a variant excludes self-medication. Case fatality among accidental poisoning admissions was 0% under 20 years and 3.0% at 20–59 years; weighted by the age of therapeutic errors, nearly half in children,^23^ it is 1.1%. Errors managed without admission add emergency care (76% of calls to the centre come from health facilities), two days of moderate acute disability for those seen in a facility, sequelae among admitted survivors and carers’ time for children. The same Δ and *s*_*F*_ then apply. A controlled-medicines scenario adds an illustrative 10% increase without a pharmacist to intentional poisonings involving them (21%; 5,734 admissions in 2023, held constant). Antibiotic courses sold without prescription are counted, but their health effects enter only an exploratory analysis.

### Valuation of health

Health lost is valued as

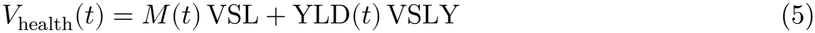

where *M* is deaths and YLD the years lived with disability, from Global Burden of Disease disability weights.^25^ Health losses are also summarized as DALYs. The value of a statistical life-year, VSLY, is the value of a statistical life (VSL) divided by 42.8 remaining years of life expectancy at mean adult age. An ADR admission costs CLP 4.14 million.

### Access and viability

Of the 300,000 residents of municipalities without a pharmacy, we assumed that 5% (range 1–15%) would gain one under the proposal, phased in over three years. Sixty per cent of those people (range 30–90%) already obtained medicines elsewhere and save time and fares; the rest had unmet need and gain health through better adherence:^26^

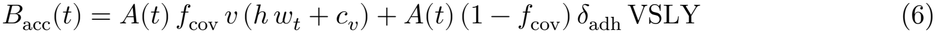

Here *A*(*t*) is people gaining a pharmacy and *f*_cov_ the share already covered. Each person makes *v* = 8 purchases a year, each saving *h* = 1.5 hours of travel, valued at the official social value of travel time, *w*_*t*_ = CLP 10,047 per hour,^14^ and a fare, *c*_V_ = CLP 3,000 (authors’ assumptions except *w*_*t*_; Supplementary Table 8). Those with unmet need gain *δ*_adh_ = 0.0005 DALYs per person-year. Separately, an exploratory viability model estimated the minimum population at which each type of outlet covers its costs (Supplementary Table 10).

### Interpreting the labour assumption

Productive recovery combines three quantities: the share of displaced pharmacists who are re-employed (*r*), their pay in the new job relative to the old (*w*), and the years before they are re-employed (*a*):

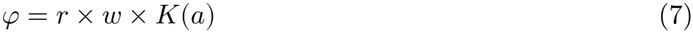

*K*(*a*) is the share of the value of their work that is kept despite the delay; because pharmacists are displaced gradually, it is computed separately for each year’s group of displaced posts (Supplementary Note 5 and Supplementary Table 18). The safety value of the pharmacist’s work is not counted here, since it is already in the health account.

### Uncertainty and statistical analysis

In total, 58 parameters were drawn in 10,000 Monte Carlo simulations from beta, gamma or lognormal distributions fitted to each declared range (seed 20261018). We report means with 95% intervals (2.5th and 97.5th percentiles of the simulations), which describe parameter uncertainty within a scenario, not the uncertainty of the scenarios themselves (Table 1). Because NPV_*c*_ is linear in Δ, *s*_*F*_ and *φ*, thresholds were obtained exactly by evaluating two values of each. Analyses used R (version 4.3.3) with the data.table, readxl and ggplot2 packages.

### Reporting standards and sex and gender

Reporting follows the Consolidated Health Economic Evaluation Reporting Standards 2022 (CHEERS 2022);^27^ the checklist is given in Supplementary Note 8. Sex and gender were not considered in the model design, because most inputs, including the scenarios that drive the results, are not available by sex; results were not disaggregated by sex or gender.

## Results

Results refer to the reference scenario unless stated, as means with 95% intervals of 10,000 simulations; monetary values are present values in billions of 2026 Chilean pesos, discounted at 3% (CLP 1 billion = US$1.1 million).

### Viability and access in municipalities without a pharmacy

Replacing the pharmacist makes small pharmacies more viable. The median minimum population an outlet needs for its sales to cover its costs was 2,420 residents for a pharmacy with a pharmacist (95% interval 1,140–4,990) and 1,350 for one with a technician (640–2,770), 44% fewer; a drugstore needs 2,120 (920–4,860). The drugstore is the cheapest outlet to run but sells only the restricted list, about three-quarters of a pharmacy’s sales (Figure 1a). Among 15 municipalities without a pharmacy listed by the national association of municipalities (2024 census), the two largest, Huara and Río Ibáñez, would gain most: the share of simulations in which a pharmacy covers its costs rises from about 65% with a pharmacist to 98% with a technician (Figure 1b). In eight, with fewer than 650 residents, no type of outlet covers its costs from sales alone.

**Figure 1.**
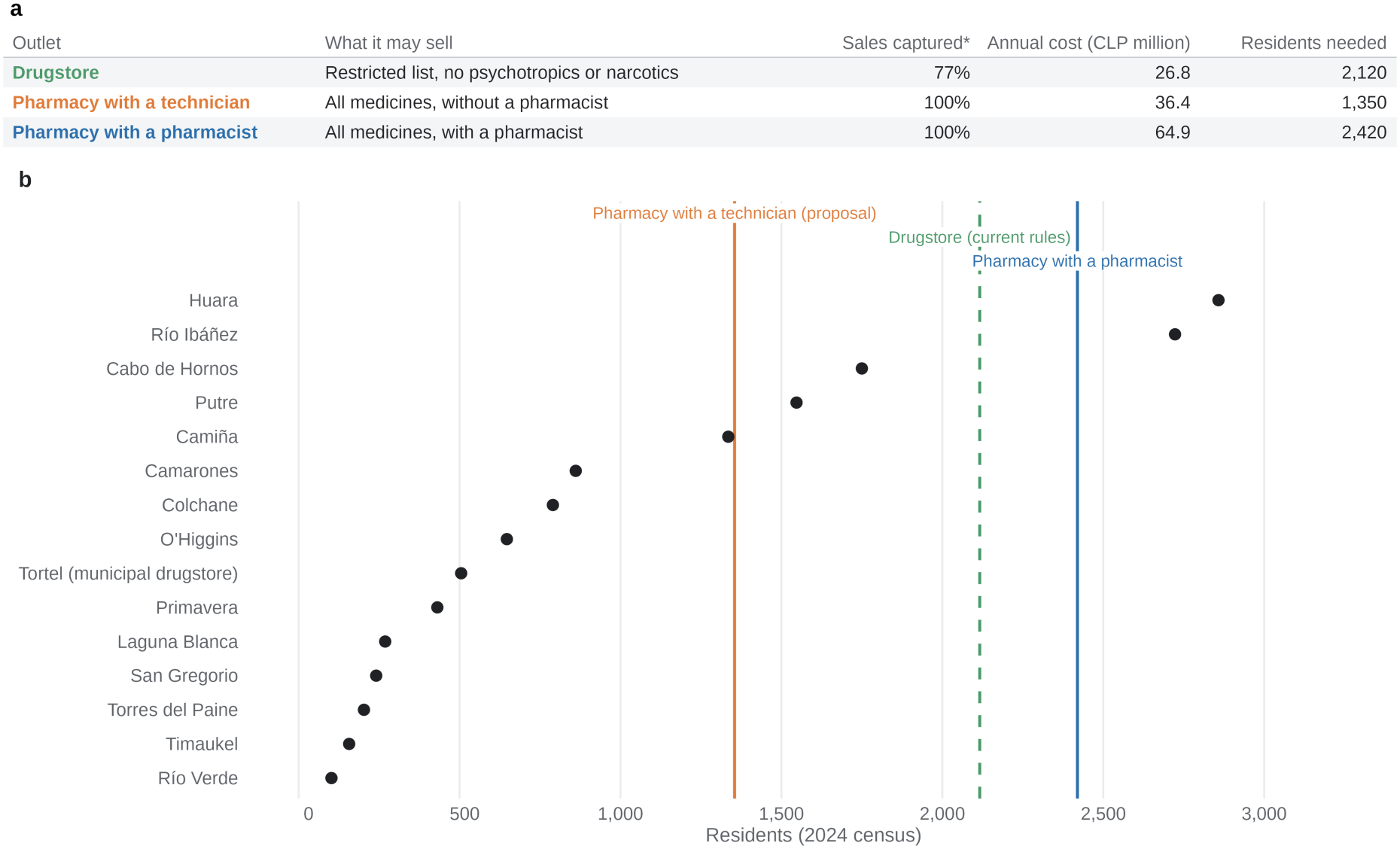
Municipalities without a pharmacy and the population an outlet needs to cover its costs. Panel a gives the medians for each type of outlet: what it may sell, the share of a pharmacy’s sales it captures (*), its annual cost (fixed costs, director, logistics and annualised investment) and the residents it needs for its sales to cover that cost. In panel b, lines show the same medians of the minimum population and points show the 2024 census population of the 15 municipalities without a pharmacy listed by the national association of municipalities. Medians of 10,000 simulations.

If 5% of residents of municipalities without a pharmacy gained one, about 17,700 people would do so from 2029 (2,800–45,400), of whom 6,500 (700–20,400) previously had unmet need. This access was worth CLP 12 billion (2–38) over ten years, about CLP 694,000 per person, nine-tenths of it in time and fares saved (Table 2).

### Exposure to dispensing without a pharmacist

With medium replacement, 2,330 full-time pharmacist posts (1,650–3,180) would be replaced by 2036, leaving 39% of opening hours nationally without a pharmacist but only 27% of dispensings, because weekend hours are replaced first (Table 2). With low and high replacement, dispensings without a pharmacist would be 8% and 54%.

### Health harm

Over ten years, the proposal would add 2,470 ADR admissions and 6,465 emergency visits, 670 poisonings from therapeutic errors (460–820), 28 of them admitted and nearly half in children, and 68 deaths (22–166), or 1,222 disability-adjusted life years (DALYs). Deaths account for 97% of the value of health lost (Table 2). Poisonings and the controlled-medicines scenario each added under one death. Deaths reached 234 with high replacement and a high increase in errors (Supplementary Table 11).

Most of the additional poisonings never reach a hospital. The national poison centre recorded 3,283 therapeutic errors in 2024, and 44.3% were of types a pharmacy could intercept, mainly wrong doses and self-medication, along with prescribing, dispensing and formulation errors (Supplementary Table 23). For each additional poisoning admission the model adds more than 20 errors managed without admission, three-quarters of them seen in a health facility. None is fatal, so they add little monetary value, but they account for nearly all the additional poisonings.

### Private savings and the net health and access value

The labour-cost savings and the net health and access value point in opposite directions (Figure 2). Owners would save CLP 270 billion (US$296 million) in pay, whereas health losses and care costs (CLP 150 billion) would be more than 12 times the access gain, and the net health and access value was negative in 99.9% of simulations. Even if pharmacies passed all the savings on to prices, annual spending on medicines would fall by only 0.6% to 1.9% (for spending of CLP 250,000 to 80,000 per person a year). With a pass-through of 30%, consumers would receive CLP 81 billion and owners would keep 189.

**Figure 2.**
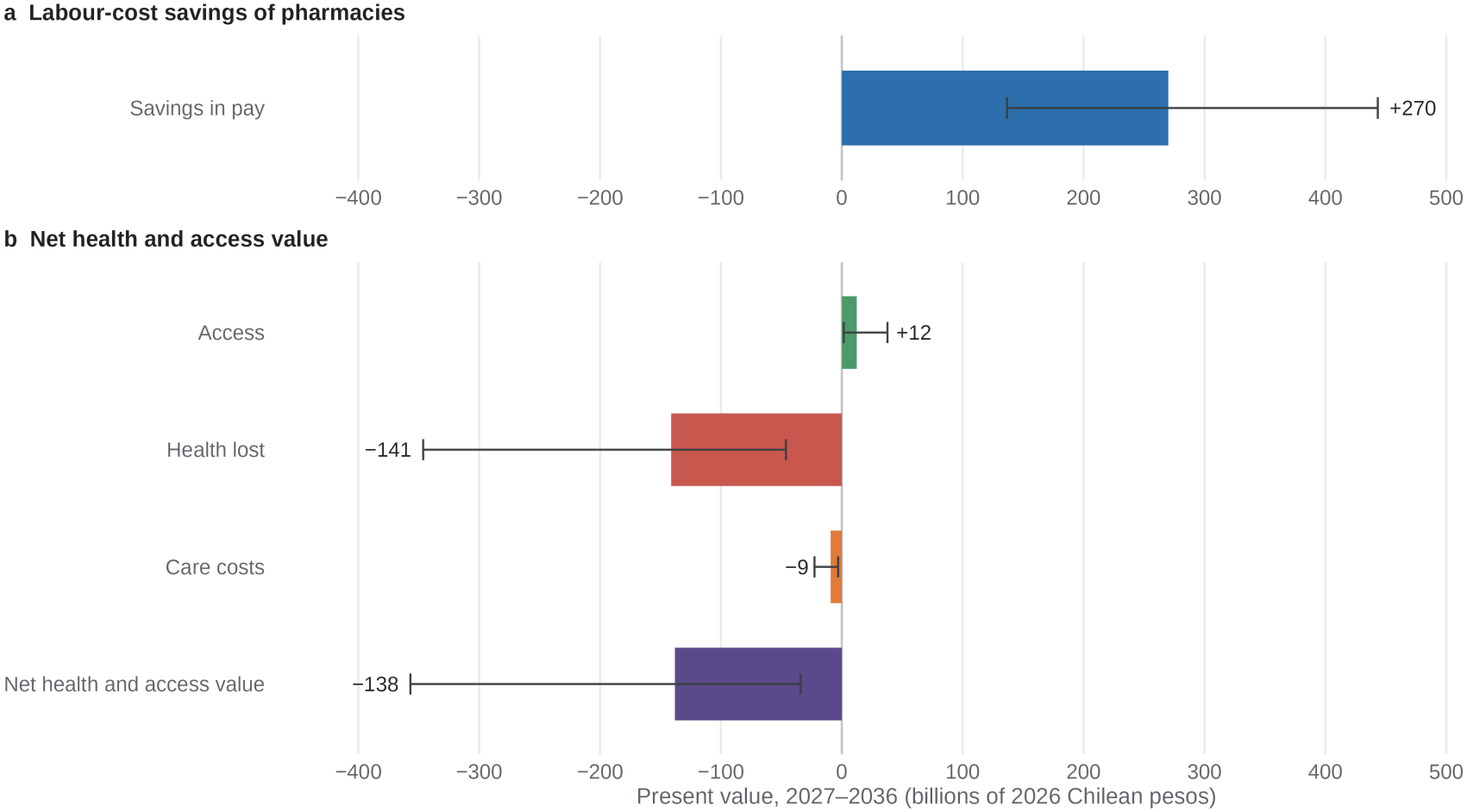
Labour-cost savings of pharmacies and the net health and access value. Panel a shows the savings in pay to pharmacy owners and panel b the components of the net health and access value, which excludes labour and price transfers. The two accounts are not added. Bars show means and error bars 95% intervals of 10,000 simulations; present values for 2027–2036 in billions of 2026 Chilean pesos, discounted at 3%, reference scenario.

### Sensitivity to unmeasured assumptions

Table 3 changes one assumption at a time, ceteris paribus, from the reference. The net health and access value was negative in every variant except that without an increase in errors, where it equals the access gain. Because it falls linearly with the increase in errors without a pharmacist, it turns negative with any increase above 3.2% (6.4% if a quarter of the harm passes through the affected pharmacies). Discounting at 5.5% or 0% changes its size but not its sign. All variants are listed in Supplementary Table 16.

**Table 3.** Effect of changing one assumption from the reference (CLP billion)

| **Change from the reference** | **Deaths over ten years** | **Net health and access value** | **Complementary social NPV with 80% recovery** | **Simulations in which the proposal is worse (NPV)** |
|---|---|---|---|---|
| None (reference) | 68 | −138 (−357 to −34) | +29 (−207 to +198) | 33% |
| No increase in errors (0%) | 0 | +12 (+2 to +38) | +179 (+76 to +312) | 0% |
| Low increase in errors (20%) | 35 | −65 (−177 to −8) | +102 (−41 to +246) | 7% |
| High increase in errors (67%) | 116 | −246 (−621 to −71) | −79 (−462 to +141) | 68% |
| 25% of harm passes through the pharmacies | 34 | −63 (−173 to −7) | +103 (−38 to +248) | 7% |
| All harm passes through the pharmacies | 135 | −288 (−723 to −85) | −121 (−561 to +120) | 76% |
| Proportional replacement across all hours | 101 | −213 (−530 to −64) | −47 (−377 to +157) | 60% |
| Case fatality of 0.5% | 13 | −27 (−83 to +10) | +140 (+30 to +276) | 0.6% |
| Case fatality of 5% | 124 | −252 (−626 to −75) | −85 (−460 to +137) | 70% |
| Case fatality and years of life lost not weighted by age (2.1%; 13.4) | 52 | −107 (−274 to −24) | +60 (−126 to +218) | 21% |
| Health valued as DALYs × VSLY | 68 | −55 (−152 to −3) | +112 (−22 to +254) | 5% |
| Controlled medicines: intentional poisonings +10% without a pharmacist | 68 | −139 (−358 to −35) | +27 (−209 to +197) | 33% |
| All unintentional poisonings, assumed 8% increase (upper bound) | 71 | −145 (−364 to −40) | +21 (−215 to +191) | 36% |
| Antibiotic resistance and adverse events included (exploratory) | 80 | −164 (−385 to −52) | +3 (−238 to +175) | 43% |
| Technician paid a 20% premium for directing the pharmacy | 68 | −138 (−357 to −34) | −21 (−257 to +146) | 54% |
| Discount rate of 5.5% | 68 | −121 (−314 to −30) | +28 (−180 to +179) | 32% |
| Discount rate of 0% | 68 | −163 (−416 to −41) | +30 (−246 to +226) | 34% |
| Baseline from 2025 discharges | 67 | −137 (−353 to −34) | +30 (−203 to +199) | 32% |
| Technician’s opportunity cost of 50% | 68 | −138 (−357 to −34) | +152 (−88 to +339) | 7% |

*Means (95% intervals) of 10,000 simulations. The 80% recovery is a comparison anchor, not an estimate, and the percentages of simulations are conditional on it. Supplementary Table 16 lists all variants. DALY, disability-adjusted life year; NPV, net present value; VSLY, value of a statistical life-year.*

### Conditions on the re-employment of displaced pharmacists

The complementary social NPV depends mainly on what replaced pharmacists produce elsewhere. It is approximately *φ* × 517 − 247 − 138 CLP billion: the work replaced pharmacists do elsewhere (a fraction *φ* of the 517 billion of labour released), minus the technicians’ previous work (247), plus the net health and access value. If replaced pharmacists produce nothing elsewhere, it is −385 billion (−630 to −226); if they produce as much as before, +132 (−118 to +338). The proposal and current regulation are equal at *φ* = 0.74, at 0.51 if the technicians’ previous work were worth half their pay (*θ* = 0.5), and at 0.84 with a 20% technician premium.

For the proposal to equal current regulation (Figure 3), 74% of displaced work must be re-employed at the same labour cost if re-employment is immediate; with one year’s delay, 85%; with two, 98%. With a common delay beyond about 2.1 years, equality is unreachable even if everyone returns at the same pay; with *θ* = 0.5 the limit is about 4.2 years.

**Figure 3.**
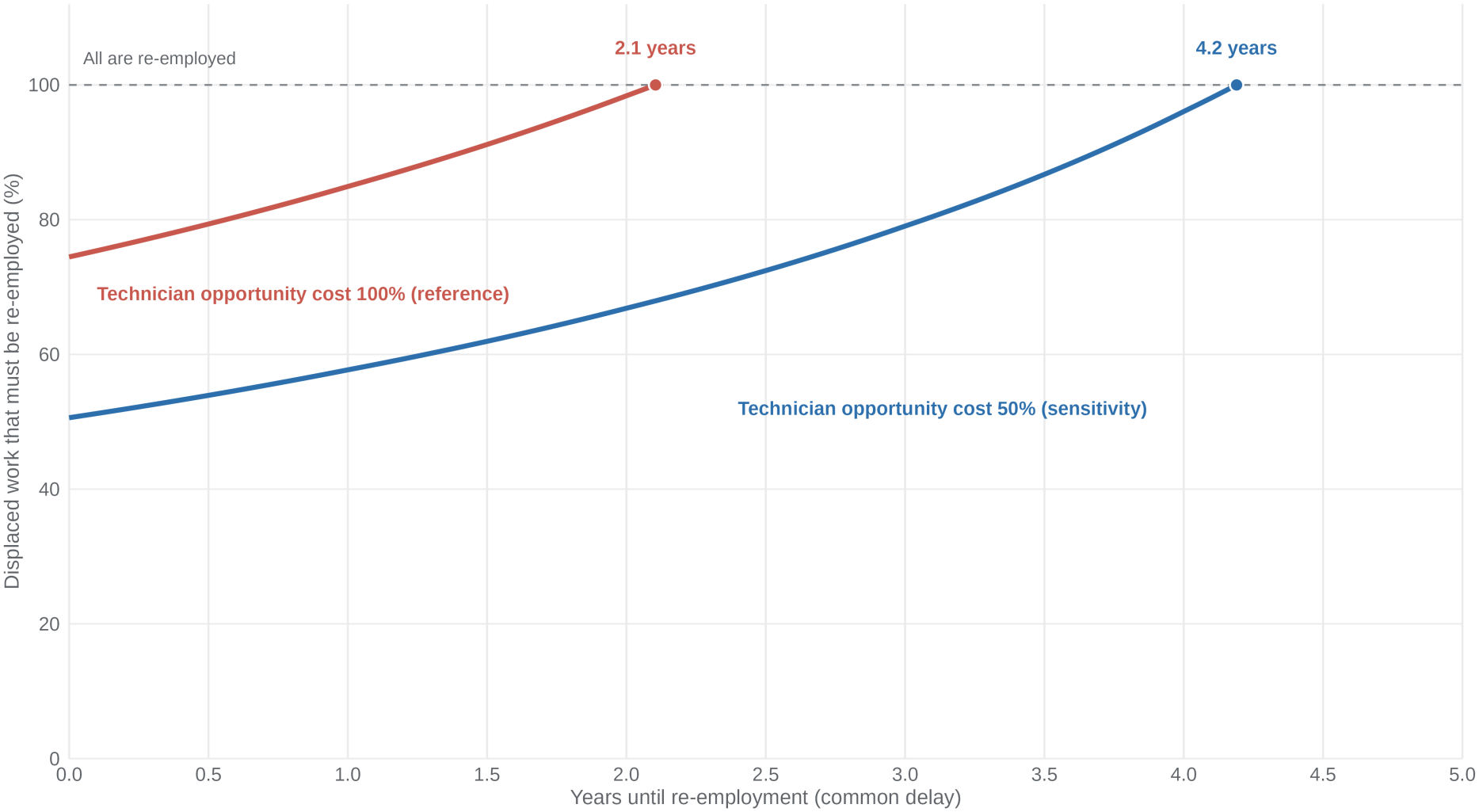
Pharmacist re-employment required to equalize the economic balance of the proposal and current regulation. Each curve marks equality of the expected complementary social net present value of the proposal and current regulation, holding clinical and access assumptions at the reference, for a technician opportunity cost of 100% (reference) or 50% of their labour cost. Above a curve the proposal has the higher value; below it, current regulation. A common re-employment delay and full recovery of the labour cost per full-time post are assumed. End points refer to the 2027–2036 horizon and shift with the health assumptions.

## Discussion

The bill rests on an access and price argument, and the model gives that argument little support. Technician-led pharmacies are more viable in municipalities of 1,000 to 3,000 residents, but where a drugstore is already allowed, they add, for about CLP 10 million more a year, a quarter of a pharmacy’s sales, drawn from medicines outside its list, including the psychotropics and narcotics kept under a pharmacist’s control. The access gain is modest (CLP 12 billion) and local, whereas replacement would be allowed in all 4,600 pharmacies, mostly chain outlets where pharmacies already exist. The links from more outlets to more competition and lower prices are not established for Chile,^6,7^ and even full pass-through of the labour savings would lower spending by less than 2%.

The mechanism of harm is plausible though its size is uncertain. Part of drug-related morbidity is preventable, part is intercepted when a prescription is checked at dispensing, and that check is where a pharmacist’s training differs most from that of non-pharmacist directors. Studies of non-prescription antibiotic sales show that a pharmacist’s presence changes what is dispensed,^19,20^ but none measures serious events, so 68 deaths is a projection under stated assumptions, not a forecast.

Poisonings added little monetary value but hundreds of events, nearly half in children under 12 years, whose doses pharmacists routinely explain. Close to half of the therapeutic errors reported to the poison centre are of the kind a pharmacy can intercept, such as wrong doses, self-medication and prescribing or dispensing errors, and the medicine most often involved is tramadol, an opioid sold on prescription.^9^

Private savings are not a social gain: the same CLP 270 billion of savings is compatible with a complementary NPV from −385 to +132 billion, depending only on what displaced pharmacists produce elsewhere. A favourable balance requires that at least 74% of displaced posts be re-employed immediately, or 98% within two years, at the same labour cost, and more than four-fifths if technicians command a premium for directing the pharmacy. Other employers would need to absorb some 2,300 posts quickly. Displaced workers often suffer persistent earnings losses,^28^ but no data exist for Chilean pharmacists, so we report no probability that the condition is met.

The question is not only Chilean. Brazil, Argentina and Peru also require a pharmacist to direct the pharmacy and be present, whereas Colombia allows drugstores to be directed by pharmacy technologists or licensed drug dispensers, and Mexico requires a health professional only where controlled medicines are sold (Supplementary Table 25). Regional experience shows that dispensing rules change what patients receive: enforcing prescription-only sales of antibiotics reduced their use in Mexico and slowed its growth in Brazil.^29^ The framework applies to proposals in the first group of countries; the second group could supply the missing evidence on errors without a pharmacist.

The study has limitations. The ADR baseline applies international rates to Chilean discharges, and case fatality comes from few deaths in coded discharges. The increase in errors without a pharmacist and the share of harm passing through pharmacies are unmeasured scenarios; the former rests on antibiotic sales abroad. Published poison-centre data do not report severity by type of error, so the same interceptable share is applied to admitted and non-admitted errors; poison-centre calls are not a sample of admissions and record no outcome after the call; the interception criteria need clinical validation; and the duration, severity and sequelae of poisonings managed without admission are assumed. Results were not disaggregated by sex or gender. Staffing and weekend workload are the authors’ judgements, and another professional as director might cost and perform differently from a technician. Access is measured only in municipalities with no pharmacy, the share of residents who gain one and most viability costs are assumed, the social NPV omits investment, training and enforcement, and parameters are drawn independently.

What decides the sign of the results could be measured before legislating or during phased implementation: how error detection changes without a pharmacist, ideally with simulated patients; the share of drug-related admissions originating in private pharmacies; poison-centre calls linked to hospital discharges and outcomes, recording where the medicine was dispensed and whether a pharmacist was present; a census of pharmacies, opening hours and pharmacist staffing; and the re-employment of displaced pharmacists. Alternatives aimed directly at access, such as an exception limited to isolated localities, remote supervision by a pharmacist^30^ or stronger municipal provision, deserve evaluation with the same framework.

## Conclusion

Allowing any pharmacy to replace its pharmacist would produce substantial private savings, small and concentrated access gains, at most a marginal fall in prices and a health loss exceeding the access gain under small increases in errors. An economic case would require most displaced pharmacists to be re-employed quickly at the same pay, which no Chilean evidence shows. On current evidence, access and prices do not justify replacing pharmacists nationwide.

## Data Availability

All data used in this study are publicly available. Hospital discharge data are available as open data from the Department of Health Statistics and Information of the Chilean Ministry of Health (https://deis.minsal.cl/#datosabiertos). Poison-centre data come from the published 2024 annual report of the Centro de Informacion Toxicologica of the Pontificia Universidad Catolica de Chile (CITUC) and a published conference abstract. All parameter values, their sources and all data produced in the present work are contained in the manuscript and its Supplementary material. The model code (R), parameter dictionary and simulation outputs will be made openly available in a public repository on journal publication.

## Declarations

## Author contributions

CB and DL conceived the study. CB, DL, JCR and ME designed the methods and provided data. CB and JCR collated and extracted the data. CB performed the formal analysis and wrote the original draft. CB, DL, JCR and ME reviewed and edited the manuscript. All authors had access to and verified the data, read and approved the final version of the manuscript and take responsibility for the decision to submit it for publication.

## Data availability

Hospital discharge data are publicly available from the Department of Health Statistics and Information of the Chilean Ministry of Health (https://deis.minsal.cl). Poison-centre data come from the centre’s published annual report^23^ and a published conference abstract.^9^ All parameter values and their sources are given in Supplementary Tables 2–10.

## Code availability

The model code (R), parameter dictionary and simulation outputs will be deposited in a public repository with a persistent identifier, without access restrictions, on publication of the peer-reviewed article, and are available from the corresponding author on request.

## Use of generative AI

During the preparation of this work, the authors used Claude (Anthropic; Opus 5.5 models, 2026) to proofread the English text and to review the R code. After using this tool, the authors reviewed and edited the content as needed, verified the code and all results against the model outputs, and take full responsibility for the content of the article.

