## Supplementary material for "Projected effects of removing pharmacists from pharmacies on access, prices and medication safety in Chile"

#### Contents

Supplementary Note 1. Model structure and equations

Supplementary Note 2. Worked example with base values

Supplementary Note 3. Parameters

Supplementary Note 4. Full results

Supplementary Note 5. Interpreting productive recovery

Supplementary Note 6. Selection of hospital discharges

Supplementary Note 7. Pharmacy direction requirements in selected Latin American countries

Supplementary Note 8. CHEERS 2022 checklist

Supplementary references

#### Supplementary Note 1. Model structure and equations

The model is an annual simulation from $t = 1$ (2027) to $t = 10$ (2036), evaluated in each of 10,000 Monte Carlo draws. The subscript $k$ denotes chains and franchises ($c$) or independent pharmacies ($i$). Supplementary Fig. 1 shows how the modules connect.

### Supplementary Fig. 1. Model structure

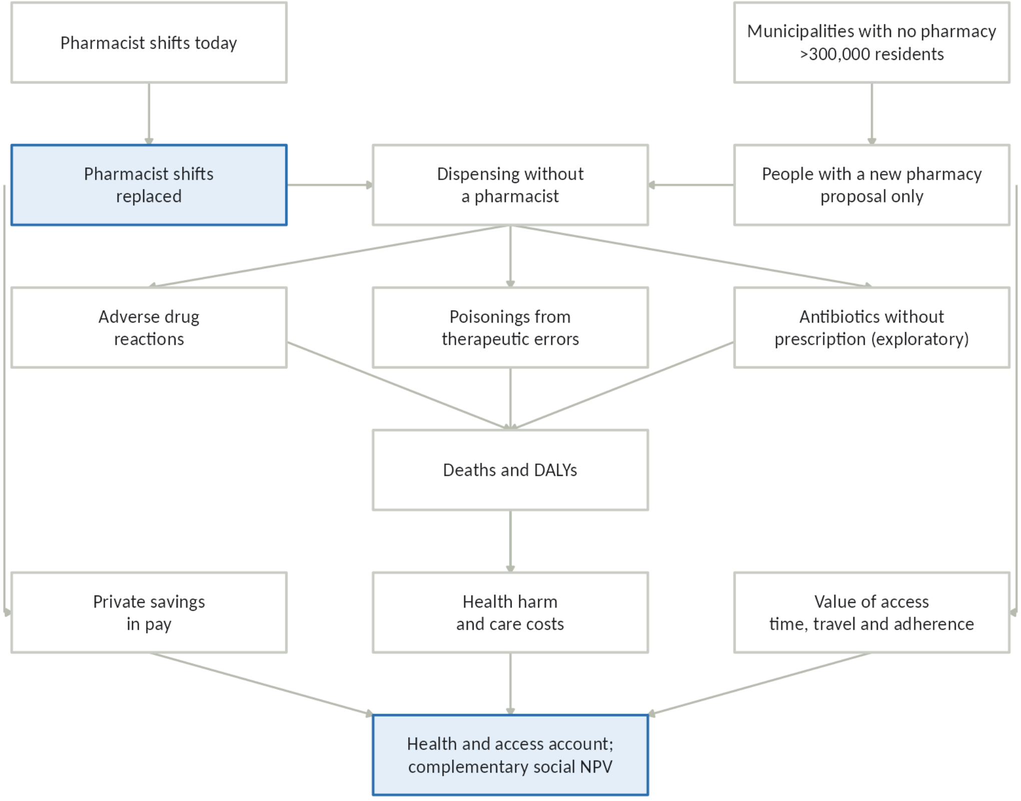


*Source: own elaboration.*

#### Pharmacist posts and replacement

Full-time pharmacist posts by type of pharmacy, $J_k$, are the number of outlets of that type times weekly opening hours, divided by the legal working week $L(t)$ (42 hours in 2027, a day-weighted 40.63 in 2028 and 40 from 2029, Law 21,561) and increased by 6% for holidays and absences. One pharmacist is present per opening hour. The share replaced, $S_k(t)$, approaches the long-run scenario level $S_k^{\max}$, reaching a fraction $a_1$ of it in the first year. Among independents only the fraction $1-d$ not directed by an owner-pharmacist can replace.

$$J_k(t) = N\,P_k\,\frac{H_k}{L(t)}\,1.06 \qquad S_k(t) = S_k^{\max}[1-(1-a_1)^t] \qquad R(t) = J_c(t)\,S_c(t) + J_i(t)\,(1-d)\,S_i(t) \tag{S1}$$

#### Dispensings without a pharmacist

A weekend hour dispenses a fraction $r_{\text{ext}}$ of a weekday hour, and the weekend is a fraction $s_{\text{ext}}$ of opening hours. The intensities of a weekday hour, $w_{\text{wd}}$, and a weekend hour, $w_{\text{we}}$, keep the mean intensity of all hours at 1. Replacing a share $S$ of hours, weekend first, leaves a share $\rho(S)$ of dispensings without a pharmacist; under the proportional rule $\rho(S) = S$.

$$w_{\mathrm{wd}} = \frac{1}{(1 - s_{\mathrm{ext}}) + r_{\mathrm{ext}}\, s_{\mathrm{ext}}} \qquad w_{\mathrm{we}} = r_{\mathrm{ext}}\, w_{\mathrm{wd}} \qquad \rho(S) = w_{\mathrm{we}} \min(S, s_{\mathrm{ext}}) + w_{\mathrm{wd}} \max(0, S - s_{\mathrm{ext}}) \tag{S2}$$

$$E_F(t) = D_c\, \rho(S_c(t)) + (1 - D_c)(1 - d)\, \rho(S_i(t)) \qquad D_N(t) = \frac{u_{\mathrm{rel}}\, A(t)}{\mathrm{Population}} \tag{S3}$$

$D_c$ is the chains' share of dispensings, $A(t)$ the people gaining a pharmacy (equation S10) and $u_{\mathrm{rel}}$ their medicine use relative to the national mean.

#### Time profile of the clinical effect

$$p(t) = g_0 + (1 - g_0) \min\left(1, \frac{t-1}{T-1}\right) \tag{S4}$$

#### Adverse drug reactions

The baseline $H_0$ is the sum over three age groups of 2019 non-surgical discharges (excluding chapters O and P and codes Z33–Z39), projected to 2026 with United Nations population ratios, times the age-specific rate of admissions associated with adverse drug reactions. Rates are drawn from a split lognormal distribution that reproduces the median and both quartiles reported by Kongkaew et al., using the same quantile for all age groups in each draw, with a floor of 2,500 admissions. The increase in errors, $\Delta$, is derived from an odds ratio OR and a baseline probability $p_0$ with a pharmacist present; $r_n = 1$ counts dispensings by new outlets as made without a pharmacist.

$$1 + \Delta = \frac{1}{p_0 + \mathrm{OR}\,(1 - p_0)} \tag{S5}$$

$$\Delta H_{\mathrm{ADR}}(t) = H_0 (1+g)^t\, f_{\mathrm{prev}}\, f_{\mathrm{disp}}\, \Delta\, p(t)\, [s_F\, E_F(t) + r_n\, D_N(t)] \tag{S6}$$

$$U(t) = r_{\mathrm{urg}}\, \Delta H_{\mathrm{ADR}}(t) \qquad M_{\mathrm{ADR}}(t) = m_{\mathrm{ADR}}\, \Delta H_{\mathrm{ADR}}(t) \tag{S7a}$$

$$\mathrm{Sec}(t) = f_{\mathrm{sec}}\, [\Delta H_{\mathrm{ADR}}(t) - M_{\mathrm{ADR}}(t)] \tag{S7b}$$

#### Poisonings and antibiotics

The poisoning baseline $H_{0,\mathrm{tox}}$ is accidental medicine poisoning (codes T36–T50 with external cause X40–X44). A fraction $f_{\mathrm{TE}}$ of unintentional poisonings are therapeutic errors. A fraction $q_{\mathrm{TE}}$ of therapeutic errors could be intercepted at the pharmacy (Supplementary Table 23); because published data do not report severity by type of error, the same fraction applies to admissions and to errors managed without admission. The same $\Delta$ and $s_F$ as in equation S6 apply. In the controlled-medicines variant, $H_{0,\mathrm{int}}\, f_{\mathrm{ctrl}}\, \Delta_{\mathrm{ctrl}}\, p(t)\, \{s_F\, E_F(t) + r_n D_N(t)\}$ intentional poisonings are added, with case fatality $m_{\mathrm{int}}$; $H_{0,\mathrm{int}}$ is held at its 2023 level rather than extrapolating its recent rise. Antibiotic courses sold without prescription, $Q(t)$, follow from the probability of sale without a pharmacist, $p_1$, obtained by dividing the odds with a pharmacist by OR.

$$\Delta H_{\text{tox}}(t) = H_{0,\text{tox}}(1+g)^t\, f_{\text{TE}}\, q_{\text{TE}}\, \Delta p(t)\, [s_F\, E_F(t) + r_n D_N(t)] \qquad M_{\text{tox}}(t) = m_{\text{tox}}\, \Delta H_{\text{tox}}(t) \quad \text{(S8a)}$$

$$Q(t) = \text{Sol}\,[E_F(t) + r_n D_N(t)]\,(p_1 - p_0)\, p(t) \quad \text{(S8b)}$$

Poisonings managed without admission are counted as well (Supplementary Note 6, Supplementary Table 24). The same chain is applied to the therapeutic errors managed without admission, $H_{0,\text{amb}}$, of which a fraction $f_{\text{urg}}$ is seen in a health facility; assigns to each of them a loss of $\text{YLD}_{\text{case}}$ years lived with disability; adds sequelae in a fraction $f_{\text{sec,tox}}$ of admitted survivors; and counts the workdays lost by carers of children under 12 years, a fraction $f_{\text{ch}}$ of therapeutic errors. No deaths are added outside hospital.

$$\Delta A_{\text{tox}}(t) = H_{0,\text{amb}}(1+g)^t\, q_{\text{TE}}\, \Delta p(t)\, [s_F\, E_F(t) + r_n D_N(t)] \qquad U_{\text{tox}}(t) = f_{\text{urg}}\, \Delta A_{\text{tox}}(t) \quad \text{(S8c)}$$

$$\text{YLD}_{\text{amb}}(t) = \Delta A_{\text{tox}}\, \text{YLD}_{\text{case}} + f_{\text{sec,tox}}(\Delta H_{\text{tox}} - M_{\text{tox}})\, dw_{\text{sec}}\, dur_{\text{sec}} \quad \text{(S8d)}$$

$$C_{\text{amb}}(t) = U_{\text{tox}}\, c_{\text{urg}} + f_{\text{ch}}\, (\Delta H_{\text{tox}}\, d_{\text{hosp}} + U_{\text{tox}}\, d_{\text{amb}})\, 8\, w_t \quad \text{(S8e)}$$

$\text{YLD}_{\text{amb}}$ is added to YLD (equation S9b) and valued at the VSLY; $C_{\text{amb}}$, which includes carers' time, is added to care costs (equation S9e).

In the exploratory analysis, resistance deaths in year $t$ are $M_{\text{AMR}}\, \varepsilon\, w_{\text{comm}}\, Q(t-1)/Q_{\text{total}}$. Each is accompanied by resistant infections and their excess cost, and each course generates emergency visits for adverse events (9.2% admitted).

#### DALYs, valuation and care costs

$$\text{YLD}_{\text{ADR}}(t) = (\Delta H_{\text{ADR}} - M_{\text{ADR}})\, dw_{\text{ADR}} \frac{dur_{\text{ADR}}}{365} + U\, dw_{\text{urg}} \frac{dur_{\text{urg}}}{365} + \text{Sec}\, dw_{\text{sec}}\, dur_{\text{sec}} \quad \text{(S9a)}$$

$$\text{YLD}(t) = \text{YLD}_{\text{ADR}}(t) + (\Delta H_{\text{tox}} - M_{\text{tox}})\, dw_{\text{tox}} \frac{dur_{\text{tox}}}{365} \quad \text{(S9b)}$$

$$\text{YLL}(t) = M_{\text{ADR}}\, \text{YLL}_{\text{ADR}} + M_{\text{tox}}\, \text{YLL}_{\text{tox}} \quad \text{(S9c)}$$

$$V_{\text{health}}(t) = (M_{\text{ADR}} + M_{\text{tox}})\, \text{VSL} + \text{YLD}(t)\, \text{VSLY} \quad \text{(S9d)}$$

$$C_{\text{care}}(t) = \Delta H_{\text{ADR}}\, c_{\text{ADR}} + U\, c_{\text{urg}} + \Delta H_{\text{tox}}\, c_{\text{tox}} \quad \text{(S9e)}$$

$\text{VSLY} = \text{VSL}/42.82$. In the age-adjusted variant, $V_{\text{health}} = (\text{YLL}+\text{YLD})\,\text{VSLY}$. In the transferred-VSL variant, $\text{VSL} = \text{VSL}_{\text{US}}\,(\text{GNI}_{\text{Chile}}/\text{GNI}_{\text{US}})^e\,\text{PPP}$ with income elasticity $e$ of 0.8 or 1.

#### Access

$$A(t) = \mathrm{Pop}_{\mathrm{none}}\, p_{\mathrm{ent}} \min(1, t/3) \qquad B_{\mathrm{acc}}(t) = A(t)\, f_{\mathrm{cov}}\, v\, (h\, w_t + c_v) + A(t)\, (1 - f_{\mathrm{cov}})\, \delta_{\mathrm{adh}}\, \mathrm{VSLY} \quad \text{(S10)}$$

#### Viability of a small outlet

The population at which an outlet covers its costs, $N_{\mathrm{min}}$, is its annual cost divided by the net margin left by each resident. Costs are monthly fixed costs $F$, logistics $L_{\mathrm{og}}$ and director's pay $D$ with employer contributions $c_p$ and staff cover $k$, plus the initial investment $I$ annualised with the capital recovery factor FRC (ten years at 5.5%, a private cost of capital rather than the social discount rate). The margin is spending captured per resident $s$ times gross margin $m$, less the financing cost $r_k$ of $\mu$ months of inventory.

$$N_{\mathrm{min}} = \frac{12\,[F + L_{\mathrm{og}} + D\,(1 + c_p)\,k] + I \cdot \mathrm{FRC}}{s\,m - s\,(1 - m)\,\frac{\mu}{12}\,r_k} \quad \text{(S11)}$$

Inputs are in Supplementary Table 10. The pharmacy gross margin, 30% to 40%, brackets the 36% to 41% reported for the chains [1] and the 40% assumed in a business plan for an independent pharmacy [2]; the same plan puts fit-out and systems at about CLP 14 million, which informs the pharmacy investment, with inventory costed separately. The drugstore captures 60% to 90% of a pharmacy's spending because its authorised list covers the common medicines for chronic disease and the usual antibiotics [3]; the first municipal drugstore opened with 47 medicines [4]. Fixed costs, logistics, inventory and staff cover remain the authors' assumptions. The median spending captured per resident, about CLP 80,000 a year, equals national spending on medicines divided by the population; it is the input that matters most (Spearman rank correlation with $N_{\mathrm{min}}$ of $-0.93$ for pharmacies directed by a pharmacist or a technician, against 0.24 and 0.10 for the director's pay).

The median annual cost of running each outlet, the numerator of equation S11, is CLP 64.9 million (95% interval 52.1 to 80.4) for a pharmacy with a pharmacist, 36.4 million (29.6 to 44.4) with a technician and 26.8 million (21.3 to 33.0) for a drugstore. A pharmacy directed by a technician costs about CLP 9.7 million (1.3 to 18.5) a year more than a drugstore; for that difference it sells the medicines outside the drugstore list, including psychotropics and narcotics. Replacing the pharmacist saves a further CLP 28.2 million (16.9 to 42.7) a year.

The model counts only the director and no other staff, which understates the costs of every outlet and overstates how much replacing the pharmacist helps. With one assistant (CLP 0.5 to 0.9 million a month) in every outlet, the median $N_{\mathrm{min}}$ rises to 2,840 residents with a pharmacist, 1,780 with a technician and 3,010 for a drugstore (whose smaller margin per resident makes an extra salary weigh more), and the reduction from replacing the pharmacist falls from 44% to 37%; the population of the 15 municipalities living where only a technician pharmacy is viable stays at 4,632 in the median simulation.

Since 2020, Law 21,198 allows private pharmacies and drugstores to buy through Cenabast [5]. If pharmacies sold close to those purchase prices, as drugstores do, their margin would fall to the drugstore's range (15% to 30%): the median $N_{\mathrm{min}}$ would rise to 4,030 with a pharmacist and 2,270 with a technician, against 2,120 for a drugstore, and the reduction from replacing the pharmacist would stay at 44%. Public pharmacies buying through Cenabast sold identical medicines 64% to 68% cheaper than private ones [6], but they sell at or near cost, so that gap shows the scope for lower prices rather than the margin a private outlet would keep. Cenabast access lowers costs for pharmacies with and without a pharmacist alike and does not change the comparison between them.

#### Labour, accounts and distribution

$$C_{\mathrm{QF}}(t) = R(t)\,W_{\mathrm{QF}}\,12\,[1 + c(t)] \qquad C_T(t) = R(t)\,W_T\,12\,[1 + c(t)] \tag{S12}$$

Employer contributions $c(t)$ add unemployment insurance (2.4%), occupational accident insurance (0.93%), the childcare insurance (0.03%) and the new employer pension contribution (Law 21,735), which is 3.5% from August 2026 and rises each August to 8.5% in 2033 [7]; averaged by calendar year it rises from 3.81% in 2027 to 8.5% from 2034. The accounts are given in equations 1 and 2 of the main text; the decomposition of productive recovery is equation 7. The complementary social NPV can be split among groups, and the parts add up exactly to it:

$$\mathrm{NPV}_c = (1-\pi)\,\mathrm{PS} + \pi\,\mathrm{PS} - (1-\varphi)\,\mathrm{PV}(C_{\mathrm{QF}}) + (1-\theta)\,\mathrm{PV}(C_T) + \mathrm{NHA} \tag{S13}$$

The terms are, in order, the gains of owners and of consumers ($\pi$ is the share of savings passed on to prices), the loss of replaced pharmacists, the gain of technicians, and the net health and access value, which itself splits into health lost by the population, care costs borne by the health system and access gained by residents of municipalities without a pharmacy.

#### Decomposition of productive recovery

Posts released at the start of year $s$, $\Delta R(s) = R(s) - R(s-1)$, return to productive work after a common delay $a$. In year $t$ they contribute the fraction $\min\{1, \max(0, t - s + 1 - a)\}$ of their labour cost in that year. Summing over cohorts in each draw,

$$K(a) = \frac{E[\sum_t \sum_{s \le t} \Delta R(s)\, \frac{C_{\mathrm{QF}}(t)}{R(t)}\, \min[1, \max(0, t - s + 1 - a)]\, 1.03^{-(t-1)}]}{E[\mathrm{PV}(C_{\mathrm{QF}})]} \tag{S14a}$$

$$r\,w = \frac{\theta\, E[\mathrm{PV}(C_T)] - E[\mathrm{NHA}]}{K(a)\, E[\mathrm{PV}(C_{\mathrm{QF}})]} \tag{S14b}$$

Equation S14b gives the combinations of re-employed share $r$ and relative labour cost $w$ at which the expected complementary NPV of the proposal equals that of current regulation.

#### Uncertainty

Beta distributions are used for proportions, gamma for counts, costs and durations, and lognormal for ratios, each fitted so that its 2.5th and 97.5th percentiles match the declared interval. Apart from the age groups of the baseline burden, parameters are drawn independently. All scenarios use the same 10,000 draws (seed 20261018), so differences between scenarios are not due to sampling.

### Supplementary Note 2. Worked example with base values

Supplementary Table 1 evaluates the model once with the base value of every parameter and the reference scenarios, for 2036, so that each step can be reproduced by hand. Reported results are means over 10,000 draws and differ slightly from this single evaluation; for example, ten-year ADR admissions are 2,486 here and 2,470 on average.

**Supplementary Table 1. Step-by-step calculation for 2036 with base values (medium replacement, weekend first, $\Delta = 0.39$, $s_F = 0.50$)**

| Step | Calculation | Result |
|---|---|---|
| Pharmacist posts needed, chains | 4,600 × 0.54 × 75/40 × 1.06 | 4,937 |
| Pharmacist posts needed, independents | 4,600 × 0.46 × 60/40 × 1.06 | 3,364 |
| Share replaced, chains; independents | $0.45 \times (1 - 0.5^{10})$; $0.30 \times (1 - 0.5^{10})$ | 0.4496; 0.2997 |
| Posts replaced | 4,937 × 0.4496 + 3,364 × 0.18 × 0.2997 | 2,401 |
| Hour intensities | $w_{\text{wd}} = 1/(0.73 + 0.30 \times 0.27)$; $w_{\text{we}} = 0.30 \times 1.233$ | 1.233; 0.370 |
| Dispensings without a pharmacist, chains | 0.370 × 0.27 + 1.233 × (0.4496 − 0.27) | 0.321 |
| Dispensings without a pharmacist, all existing pharmacies | 0.85 × 0.321 + 0.15 × 0.18 × 0.137 | 0.277 |
| Exposure from new outlets | 0.8 × 15,000 / 20,150,948 | 0.0006 |
| ADR baseline in 2036 | $67{,}695 \times 1.0174^{10}$ | 80,440 |
| Preventable ADR admissions | 80,440 × 0.52 | 41,829 |
| Interceptable at dispensing | 41,829 × 0.15 | 6,274 |
| Extra errors reaching patients if no dispensing had a pharmacist | 6,274 × 0.39 | 2,447 |
| Exposed share: affected pharmacies plus new outlets | 0.50 × 0.277 + 0.0006 | 0.139 |
| Additional ADR admissions ($p = 1$ in 2036) | 2,447 × 0.139 | 340 |
| Emergency visits; deaths; sequelae | 2.66 × 340; 0.0274 × 340; 0.10 × (340 − 9.3) | 905; 9.3; 33.1 |
| Poisoning baseline in 2036; interceptable therapeutic errors that lead to admission | $670 \times 1.0174^{10}$; 796 × 0.208 × 0.443 | 796; 73.4 |
| Additional poisoning admissions; deaths | 73.4 × 0.39 × 0.139; × 0.0113 | 4.0; 0.04 |
| Health lost | 9.36 deaths × CLP 2,384 million + 13.1 YLD × CLP 55.7 million | CLP 23.0 billion |
| Care costs | 340 × 4.14 million + 905 × 103,400 + 4.0 × 3.10 million | CLP 1.5 billion |
| Access benefit (from 2029) | 9,000 × 8 × (1.5 × 10,047 + 3,000) + 6,000 × 0.0005 × 55.7 million | CLP 1.47 billion |
| Labour cost released; technicians hired | 2,401 × 2.2 million × 12 × 1.1186; 2,401 × 1.05 million × 12 × 1.1186 | CLP 70.9 billion; CLP 33.8 billion |
| Private savings in 2036 | 70.9 − 33.8 | CLP 37.1 billion |

### Supplementary Note 3. Parameters

Supplementary Tables 2 to 10 present each parameter with its base value, its distribution, the 95% interval to which it was fitted and the origin of that interval. The second-to-last column shows the median and mean actually implemented, which may differ from the base value because each distribution is fitted to the stated percentiles. "Observed value ±20%" denotes a measured value with no published interval; "Assumption ±50%" denotes a value with no direct source, which is the first worth measuring; "Constructed assumption" denotes a value the author derived by combining published studies and which does not correspond to a published confidence interval.

**Supplementary Table 2. Pharmacy market**

| Parameter | Base | Distribution | 95% interval | Interval origin | Median / mean in the model | Source |
|---|---|---|---|---|---|---|
| Pharmacies in operation ($N$) | 4,600 | Gamma | 3,680 to 5,520 | Observed value ±20% | 4,538 / 4,554 | FNE EM03-2018 [1] (3,809 outlets in 2015-2018); Colliers [8] |
| Share of outlets belonging to chains and franchises ($P_c$) | 0.54 | Beta | 0.432 to 0.648 | Observed value ±20% | 0.541 / 0.541 | FNE [1] (42% in 3 chains); Colliers [8] (Metropolitan Region: 49%) |
| Share of dispensings in chains and franchises ($D_c$) | 0.85 | Beta | 0.8 to 0.9 | Range across studies | 0.855 / 0.854 | FNE [1] (80% of sales in 3 chains) plus franchises |
| Weekly opening hours, chain ($H_c$) | 75 | Gamma | 60 to 90 | Observed value ±20% | 74.0 / 74.3 | Monday to Friday 9:00 to 20:00 and about 10 h on Saturday and 10 h on Sunday |
| Weekly opening hours, independent ($H_i$) | 60 | Gamma | 48 to 72 | Observed value ±20% | 59.2 / 59.4 | Typical hours of a neighbourhood pharmacy |
| Fraction of opening hours at the weekend ($s_{\text{ext}}$) | 0.27 | Beta | 0.22 to 0.32 | Defined range | 0.268 / 0.269 | Chain: Monday to Friday 9:00 to 20:00 (55 h) and about 10 h on Saturday and on Sunday (20 h); almost no pharmacy opens at night |
| Pharmacists present at the same time per outlet | 1 | Fixed | — | Fixed | — | Author's operational knowledge, pending validation |
| Mark-up for holidays and absences | 0.06 | Fixed | — | Fixed | — | Convention |
| Independents whose technical director is the owner pharmacist ($d$) | 0.82 | Beta | 0.75 to 0.9 | Assumption (defined range) | 0.834 / 0.832 | Authors' assumption from knowledge of the sector; one-way sensitivity from 0.60 to 0.95 changes deaths from 64 to 62 and the complementary NPV from +53 to +32 (base +39), because chains account for nine-tenths of replaced posts |

#### Supplementary Table 3. Pharmacist replacement

| Parameter | Base | Distribution | 95% interval | Interval origin | Median / mean in the model | Source |
|---|---|---|---|---|---|---|
| Pharmacist positions replaced in the long run, chain ($S_c^{\max}$) | 0.45 | Scenario | — | Scenario | — | No direct evidence: scenarios 0.25 / 0.45 / 0.70 |
| Pharmacist positions replaced in the long run, independents without an owner pharmacist ($S_i^{\max}$) | 0.3 | Scenario | — | Scenario | — | No direct evidence: scenarios 0.15 / 0.30 / 0.50 |
| Fraction of long-run replacement reached in year 1 ($a_1$) | 0.5 | Beta | 0.25 to 0.75 | Assumption ±50% | 0.5 / 0.5 | Author's assumption |
| Weekend dispensings per hour, as a fraction of a weekday hour ($r_{\text{ext}}$) | 0.3 | Beta | 0.2 to 0.4 | Expert judgement (defined range) | 0.293 / 0.295 | Judgement of the author as a community pharmacist: at weekends sales per hour fall to about 30% of a weekday (20% to 40%; varies by municipality) |
| Replacement rule (0 = weekend first, 1 = proportional across all hours) | 0 | Scenario | — | Scenario | — | Author's behavioural hypothesis; 0 in the reference and 1 as the alternative rule |
| Dispensings of new outlets counted as dispensings without a pharmacist ($r_n$; 1 = yes) | 1 | Scenario | — | Scenario | — | 1 in the reference (approximation: the previous channel is treated as if it had a pharmacist); 0 = same risk as the previous channel |

### Supplementary Table 4. Labour and distribution

| Parameter | Base | Distribution | 95% interval | Interval origin | Median / mean in the model | Source |
|---|---|---|---|---|---|---|
| Gross monthly salary of a community pharmacist (CLP 2026) ($W_{\mathrm{QF}}$) | 2.2 million | Gamma | 1.76 million to 2.64 million | Observed value ±20% | 2.17 million / 2.18 million | mifuturo.cl [9], income in the fourth year (2.2 to 2.4 million) |
| Gross monthly salary of a higher-level technician (CLP 2026) ($W_T$) | 1.05 million | Gamma | 840,000 to 1.26 million | Observed value ±20% | 1.04 million / 1.04 million | mifuturo.cl [9], Pharmacy Technician in the fourth year (0.9 to 1.1 million) |
| Include employer social security contributions (1 = yes) | 1 | Fixed | — | Fixed | — | Law 19,728, Law 16,744, Law 21,063, Law 21,735 |
| Productive recovery of the replaced pharmacist, fraction of previous labour cost ($\varphi$) | No preferred value | Scenario | — | Scenario | — | Grid 0 / 0.25 / 0.50 / 0.65 / 0.80 / 1; 0.80 used only as a comparison anchor; context in Davis and von Wachter [10] |
| Opportunity cost of the hired technician, fraction of labour cost ($\theta$) | 1 | Scenario | — | Scenario | — | Grid 0.50 / 0.75 / 1.00 |
| Pay premium of the technician for directing the pharmacy | 0 | Scenario | — | Scenario | — | Variant 20% (legal responsibility of the technical director); the ±20% salary range already reaches this value |
| Fraction of the saving passed on to prices ($\pi$) | 0 | Scenario | — | Scenario | — | Scenario (0 / 0.3 / 0.6); distributional only |

**Supplementary Table 5. Adverse drug reactions**

| Parameter | Base | Distribution | 95% interval | Interval origin | Median / mean in the model | Source |
|---|---|---|---|---|---|---|
| Admissions associated with community-acquired ADRs, 2026 level ($H_0$) | 67,695 | Simulated quantiles (split lognormal by age) | 36,471 to 145,642 | Range across studies (assumed distribution) | 67,695 / 72,894 | Kongkaew [11] by age (median and IQR reproduced) applied to 2019 non-surgical discharges (DEIS) projected to 2026 with the national population projections by age [12]; 2025 discharges in Supplementary Note 6 |
| Share of harm passing through pharmacies: fraction of baseline community ADR and therapeutic-error admissions linked to medicines dispensed in the pharmacies affected by the bill ($s_F$) | 0.50 | Scenario | — | Scenario | — | Working reference with no probability attached; grid 0.25 / 0.50 / 0.75 / 1; not a Chilean estimate or an interval. Context: Matute, Castillo-Laborde et al. [13] |
| Annual growth of the baseline (ageing) ($g$) | 0.0174 | Beta | 0.0087 to 0.0261 | Assumption ±50% | 0.016 / 0.016 | Growth in expected burden 2026-2036 due to age structure (INE, base 2024) |
| Preventable fraction ($f_{\text{prev}}$) | 0.52 | Beta | 0.42 to 0.62 | Published CI | 0.521 / 0.52 | Hakkarainen [14] (52%, 95% CI 42-62) |
| Preventable fraction that can be intercepted at dispensing ($f_{\text{disp}}$) | 0.15 | Beta | 0.075 to 0.225 | Assumption ±50% | 0.139 / 0.142 | Assumption informed by Howard [15]; no channel discount |

| Parameter | Base | Distribution | 95% interval | Interval origin | Median / mean in the model | Source |
|---|---|---|---|---|---|---|
| Relative increase in errors not intercepted without a pharmacist ($\Delta$) | 0.39 | Scenario | — | Scenario | — | Illustrative scenarios 0 / 0.20 / 0.39 / 0.67; 0.39 converts the constructed OR of 0.60 into a relative risk with an assumed probability of 0.30 of sale without prescription when a pharmacist is present (Methods, health harm) |
| Fraction of the long-run effect present in year 1 ($g_0$) | 0.5 | Beta | 0.25 to 0.75 | Assumption ±50% | 0.5 / 0.5 | Assumption informed by Gedeborg [16] |
| Years until full effect ($T$) | 5 | Gamma | 2.5 to 7.5 | Assumption ±50% | 4.55 / 4.667 | Assumption informed by Gedeborg [16] |
| In-hospital case fatality of ADR admissions, weighted by the age mix of $H_0$ ($m_{\mathrm{ADR}}$) | 0.0274 | Beta | 0.0219 to 0.0332 | Resampling interval | 0.0272 / 0.0273 | DEIS 2019 and 2023 by age group (Supplementary Table 20); comparison with Pirmohamed [17] (about 2.3%); variants 0.5% / 1% / 2.1% (unweighted) / 3% / 5% |
| Cost of an ADR admission (CLP 2026) ($c_{\mathrm{ADR}}$) | 4.14 million | Gamma | 3.31 million to 4.96 million | Observed value ±20% | 4.08 million / 4.09 million | 2025 DRG base price × mean weight 1.21 [18], updated by CPI |
| Disability weight, ADR admission ($dw_{\mathrm{ADR}}$) | 0.133 | Beta | 0.088 to 0.19 | Published CI | 0.134 / 0.135 | GBD, severe acute episode [19] |
| Duration of the episode including convalescence (days) ($dur_{\mathrm{ADR}}$) | 21 | Gamma | 10.5 to 31.5 | Assumption ±50% | 19.1 / 19.6 | Author's assumption (admission plus convalescence) |

| Parameter | Base | Distribution | 95% interval | Interval origin | Median / mean in the model | Source |
|---|---|---|---|---|---|---|
| Years of life lost per ADR death ($\text{YLL}_{\text{ADR}}$) | 16.9 | Gamma | 13.52 to 20.28 | Observed value ±20% | 16.7 / 16.7 | Age of ADR deaths, DEIS 2019 and 2023, with the WHO life table [20] for Chile, weighted as case fatality (Supplementary Table 20); 13.4 with the age distribution of Budnitz [21] |
| Emergency visits for ADR without admission per admission ($r_{\text{urg}}$) | 2.66 | Gamma | 2 to 3.3 | Range across studies | 2.596 / 2.61 | Shehab [22] (27.3% of visits end in admission) |
| Disability weight, ADR treated in the emergency department ($dw_{\text{urg}}$) | 0.051 | Beta | 0.032 to 0.074 | Published CI | 0.05 / 0.051 | GBD, moderate acute episode [19] |
| Duration of the episode treated in the emergency department (days) ($dur_{\text{urg}}$) | 5 | Gamma | 2.5 to 7.5 | Assumption ±50% | 4.55 / 4.667 | Author's assumption |
| Cost of an emergency visit for ADR (CLP 2026) ($c_{\text{urg}}$) | 103,400 | Gamma | 51,700 to 155,100 | Assumption ±50% | 94,097 / 96,521 | MAI emergency tariff plus tests (assumption) |
| Fraction of ADR admissions with sequelae ($f_{\text{sec}}$) | 0.1 | Beta | 0.05 to 0.15 | Assumption ±50% | 0.092 / 0.094 | Author's assumption; variant without sequelae |
| Disability weight of sequelae ($dw_{\text{sec}}$) | 0.1 | Beta | 0.05 to 0.15 | Assumption ±50% | 0.092 / 0.094 | Author's assumption |
| Duration of sequelae (years) ($dur_{\text{sec}}$) | 3 | Gamma | 1.5 to 4.5 | Assumption ±50% | 2.73 / 2.8 | Author's assumption |

#### Supplementary Table 6. Poisonings from therapeutic errors

| Parameter | Base | Distribution | 95% interval | Interval origin | Median / mean in the model | Source |
|---|---|---|---|---|---|---|
| Admissions for accidental medicine poisoning, T36–T50 with X40–X44 ($H_{0,\text{tox}}$) | 670 | Gamma | 536 to 804 | Observed value ±20% | 661 / 663 | DEIS: 704 (2019) and 637 (2023) (Supplementary Table 21) |
| Share of unintentional poisonings that are therapeutic errors ($f_{\text{TE}}$) | 0.208 | Beta | 0.15 to 0.27 | Defined range | 0.206 / 0.207 | Poison centre 2024: 3,283 / (12,476 + 3,283) [23] |
| Share of therapeutic errors interceptable at the pharmacy ($q_{\text{TE}}$) | 0.443 | Scenario | — | Scenario | — | Poison centre 2024: broad criterion 44.3%; excluding self-medication 24.8%; strict 3.4% (Supplementary Table 23); applied to admissions and to errors managed without admission |
| Case fatality, poisoning from therapeutic errors ($m_{\text{tox}}$) | 0.0113 | Beta | 0.0044 to 0.0201 | Resampling interval | 0.0103 / 0.0108 | DEIS case fatality by age weighted by the age of therapeutic errors in the poison centre 2024 (Supplementary Table 22); unweighted, 0.60% |
| Admissions with undetermined intent, Y10–Y14 (variant only) ($H_{0,\text{ind}}$) | 496 | Gamma | 397 to 595 | Observed value ±20% | 489 / 491 | DEIS 2019 and 2023; case fatality 0.30% |
| Additional annual growth of the poisoning baseline (variant only) | 0.028 | Scenario | — | Scenario | — | Poison-centre medication errors 2019–2024, 4.4% a year, less 1.6% ageing [24] |
| Admissions for intentional medicine poisoning, X60–X64, 2023 level held constant (variant only) ($H_{0,\text{int}}$) | 5,734 | Gamma | 4,587 to 6,881 | Observed value ±20% | 5,657 / 5,677 | DEIS: 5,734 (2023); 3,315 in 2019 |

| Parameter | Base | Distribution | 95% interval | Interval origin | Median / mean in the model | Source |
|---|---|---|---|---|---|---|
| Share of poisoning agents that are controlled medicines (variant only) ($f_{\text{ctrl}}$) | 0.21 | Fixed | — | Fixed | — | Poison centre 2024: 8,414 of 40,420 agents (benzodiazepines, Z-hypnotics, psychostimulants, strong opioids) |
| Increase in intentional poisonings with controlled medicines without a pharmacist (variant only) ($\Delta_{\text{ctrl}}$) | 0.10 | Scenario | — | Scenario | — | Author's illustrative assumption; no direct evidence |
| Case fatality, intentional poisoning ($m_{\text{int}}$) | 0.0036 | Beta | 0.003 to 0.0042 | Range observed across years | 0.0036 / 0.0036 | DEIS: 0.42% (2019) and 0.30% (2023) |
| Cost of a poisoning admission (CLP 2026) ($c_{\text{tox}}$) | 3.1 million | Gamma | 1.55 million to 4.65 million | Assumption ±50% | 2.82 million / 2.9 million | DRG (assumption) |
| Disability weight, poisoning ($dw_{\text{tox}}$) | 0.163 | Beta | 0.109 to 0.227 | Published CI | 0.162 / 0.164 | GBD [19] |
| Duration of the poisoning episode (days) ($dur_{\text{tox}}$) | 10 | Gamma | 5 to 15 | Assumption ±50% | 9.1 / 9.335 | Author's assumption |
| Years of life lost per poisoning death ($\text{YLL}_{\text{tox}}$) | 39.2 | Gamma | 19.6 to 58.8 | Assumption ±50% | 35.7 / 36.6 | Age of the 8 accidental-poisoning deaths, DEIS 2019 and 2023, with the WHO life table for Chile, weighted like case fatality (Supplementary Table 22) |
| Therapeutic errors a year managed without admission ($H_{0,\text{amb}}$) | 3,144 | Scenario | — | Scenario | — | Poison centre 2024: 3,283 therapeutic errors less about 139 admitted (670 × 0.208); a lower bound, since not all cases reach the centre; variant twice this volume |

| Parameter | Base | Distribution | 95% interval | Interval origin | Median / mean in the model | Source |
|---|---|---|---|---|---|---|
| Share of these seen in a health facility ($f_{\text{urg}}$) | 0.762 | Fixed | — | Fixed | — | Poison centre 2024: share of calls made from health facilities (not reported separately for therapeutic errors); cost of an ADR emergency visit ($c_{\text{urg}}$) |
| Years lived with disability per error managed without admission ($\text{YLD}_{\text{case}}$) | 0.000213 | Scenario | — | Scenario | — | Share seen in a health facility (0.762) × GBD disability weight of a moderate acute episode (0.051; Salomon [19]) × 2 days / 365; published data do not report the severity of these cases; variants half and double |
| Admitted survivors with sequelae ($f_{\text{sec,tox}}$) | 0.05 | Scenario | — | Scenario | — | Author's assumption; half the ADR value; same weight and duration as ADR sequelae; variant 0.10 |
| Share of therapeutic errors in children under 12 years ($f_{\text{ch}}$) | 0.456 | Fixed | — | Fixed | — | Poison centre 2024, table 10 (newborns, infants, preschool and school-age children) [23] |
| Carer workdays lost per admission and per emergency visit of a child ($d_{\text{hosp}}$, $d_{\text{amb}}$) | 3 and 0.5 | Scenario | — | Scenario | — | Author's assumption; valued at 8 hours × $w_t$; variant 5 and 1 |

### Supplementary Table 7. Antibiotics (exploratory analysis, except for courses)

| Parameter | Base | Distribution | 95% interval | Interval origin | Median / mean in the model | Source |
|---|---|---|---|---|---|---|
| Include resistance and direct antibiotic ADRs (1 = yes) | 0 | Scenario | — | Scenario | — | 0 in the reference; 1 in the exploratory analysis |
| Requests for antibiotics without a prescription per year (Sol) | 1 million | Gamma | 500,000 to 1.5 million | Assumption ±50% | 910,031 / 933,469 | Assumption bounded by consumption in private pharmacies [25] |

| Parameter | Base | Distribution | 95% interval | Interval origin | Median / mean in the model | Source |
|---|---|---|---|---|---|---|
| Probability of sale without a prescription with a pharmacist present ($p_0$) | 0.3 | Beta | 0.15 to 0.45 | Assumption ±50% | 0.284 / 0.288 | Assumption; South America 78% [26] as upper bound |
| Odds ratio of sale without a prescription with a pharmacist present (OR) | 0.6 | Lognormal | 0.5 to 0.72 | Constructed assumption | 0.6 / 0.603 | Intermediate value chosen by the author between Chang [27] (0.66; 0.56-0.78) and Zawahir [28] (0.54; 0.30-0.95); not a published confidence interval |
| Antibiotic courses per year in private pharmacies ($Q_{\text{total}}$) | 7.18 million | Gamma | 5.75 million to 8.62 million | Observed value ±20% | 7.09 million / 7.11 million | Marín [25], Table 3: Chile, private pharmacy sales (IQVIA) 2019, 68.3 million DDD (11.80 DDD per 1,000 inhabitants per day over 15.8 million inhabitants) / 9.5 DDD per course |
| Deaths attributable to antimicrobial resistance in Chile ($M_{\text{AMR}}^{\text{Chile}}$) | 2,080 | Gamma | 1,750 to 2,410 | Published CI | 2,062 / 2,067 | IHME/GRAM, Chile 2021 |
| Assumed proportionality between consumption and resistance (not an estimated elasticity) ($\varepsilon$) | 0.37 | Beta | 0.185 to 0.555 | Assumption ±50% | 0.355 / 0.359 | Assumption; Shrestha [29] reports correlations, not elasticities |
| Fraction of the resistance burden attributable to community use ($w_{\text{comm}}$) | 0.6 | Beta | 0.3 to 0.9 | Assumption ±50% | 0.64 / 0.629 | Assumption |
| Years of life lost per resistance death ($\text{YLL}_{\text{AMR}}$) | 15 | Gamma | 7.5 to 22.5 | Assumption ±50% | 13.7 / 14.0 | Assumption informed by GRAM 2019 |

| Parameter | Base | Distribution | 95% interval | Interval origin | Median / mean in the model | Source |
|---|---|---|---|---|---|---|
| Resistant infections per attributable death ($i$) | 3.5 | Gamma | 1.75 to 5.25 | Assumption ±50% | 3.185 / 3.267 | Assumption |
| Excess hospital cost per resistant infection (USD) ($c_{\text{inf}}$) | 2,300 | Gamma | 1,150 to 3,450 | Assumption ±50% | 2,093 / 2,147 | Allel et al. [30] |
| Emergency visits for adverse events per course ($u$) | 0.0006 | Lognormal | 0.00046 to 0.00097 | Range across studies | 0.00067 / 0.00068 | Geller [31] |
| Exchange rate (CLP per USD) | 951 | Fixed | — | Fixed | — | World Bank [32] |

#### Supplementary Table 8. Access

| Parameter | Base | Distribution | 95% interval | Interval origin | Median / mean in the model | Source |
|---|---|---|---|---|---|---|
| Residents of municipalities with no pharmacy ($\text{Pop}_{\text{none}}$) | 300,000 | Gamma | 240,000 to 360,000 | Observed value ±20% | 295,955 / 297,008 | Cenabast [33] (55 municipalities, more than 300,000 residents) |
| Fraction of that population that gains a pharmacy in its municipality ($p_{\text{ent}}$) | 0.05 | Beta | 0.01 to 0.15 | Assumption ±50% | 0.053 / 0.06 | Assumption; see Methods, access and viability |
| Fraction of demand already covered by primary care, drugstores and delivery ($f_{\text{cov}}$) | 0.6 | Beta | 0.3 to 0.9 | Assumption ±50% | 0.64 / 0.629 | Author's assumption; defines who saves time and transport |
| Dispensings per person in municipalities without a pharmacy, relative to the national average ($u_{\text{rel}}$) | 0.8 | Gamma | 0.4 to 1.2 | Assumption ±50% | 0.728 / 0.747 | Author's assumption |
| Medicine purchases per person per year ($v$) | 8 | Gamma | 4 to 12 | Assumption ±50% | 7.28 / 7.468 | Author's assumption |
| Hours saved per visit ($h$) | 1.5 | Gamma | 0.75 to 2.25 | Assumption ±50% | 1.365 / 1.4 | Author's assumption |

| Parameter | Base | Distribution | 95% interval | Interval origin | Median / mean in the model | Source |
|---|---|---|---|---|---|---|
| Social value of inter-urban travel time (CLP/hour) ($w_t$) | 10,047 | Fixed | — | Fixed | — | MDSF, Social Prices [34] |
| Transport cost per visit (CLP 2026) ($c_v$) | 3,000 | Gamma | 1,500 to 4,500 | Assumption ±50% | 2,730 / 2,800 | Author's assumption |
| DALYs averted per resident with a previously unmet need, per year ($\delta_{\text{adh}}$) | 0.0005 | Gamma | 0.00025 to 0.00075 | Assumption ±50% | 0.00046 / 0.00047 | Assumption informed by Qato [35]; variants 0 / 0.00025 / 0.00075 |
| Fraction of new access that drugstores under the current model would already achieve ($f_{\text{alm}}$) | 0 | Scenario | — | Scenario | — | Scenario (0 / 0.5); 0 attributes all new access to the proposal |
| Population of Chile | 20.15 million | Fixed | — | Fixed | — | INE [12], 2026 projection based on the 2024 Census |

### Supplementary Table 9. Valuation

| Parameter | Base | Distribution | 95% interval | Interval origin | Median / mean in the model | Source |
|---|---|---|---|---|---|---|
| Value of a statistical life, MDSF (CLP 2026) (VSL) | 2,383.68 million | Fixed | — | Fixed | — | MDSF, Social Prices [34] (60,000 UF) |
| US VSL (USD 2025) ($\text{VSL}_{\text{US}}$) | 14.1 million | Fixed | — | Fixed | — | HHS/ASPE [36] |
| GNI per capita PPP, Chile ($\text{GNI}_{\text{Chile}}$) | 35,730 | Fixed | — | Fixed | — | World Bank |
| GNI per capita PPP, United States ($\text{GNI}_{\text{US}}$) | 89,490 | Fixed | — | Fixed | — | World Bank |
| PPP conversion factor, Chile (CLP per PPP USD) (PPP) | 453.19 | Fixed | — | Fixed | — | World Bank [32] |
| Remaining life expectancy at mean adult age | 42.82 | Fixed | — | Fixed | — | WHO, life tables for Chile; Robinson [37] |

| Parameter | Base | Distribution | 95% interval | Interval origin | Median / mean in the model | Source |
|---|---|---|---|---|---|---|
| Discount rate, costs and outcomes | 0.03 | Fixed | — | Fixed | — | Ministry of Health, methodological guide for economic evaluation [38]; variants 0 and 0.055 (MDSF, Social Prices [34]) |
| Exchange rate (CLP per US$), for reporting only | 912 | Fixed | — | Fixed | — | Central Bank of Chile, average observed dollar January–October 2026 |

#### Supplementary Table 10. Viability model inputs (95% intervals)

| Input | Symbol | Pharmacy | Drugstore | Distribution |
|---|---|---|---|---|
| Annual spending per resident captured by the outlet | $s$ | CLP 40,000 to 160,000 | 60% to 90% of a pharmacy's | Lognormal; beta |
| Gross margin | $m$ | 30% to 40% | 15% to 30% | Beta |
| Monthly fixed costs (rent, energy, systems) | $F$ | CLP 0.6 to 1.5 million | CLP 0.3 to 0.9 million | Gamma |
| Monthly logistics (supply, losses due to distance) | $L_{\text{og}}$ | CLP 0.15 to 0.6 million | CLP 0.15 to 0.6 million | Gamma |
| Director's gross monthly pay | $D$ | Pharmacist: CLP 1.76 to 2.64 million plus a remote-area allowance of 10% to 50%; technician: CLP 0.84 to 1.26 million | Assistant: CLP 0.5 to 0.9 million | Gamma; beta |
| Employer contributions | $c_p$ | 15% | 15% | Fixed |
| Staff cover for holidays and leave | $k$ | 1.10 to 1.30 | 1.10 to 1.30 | Beta |
| Initial investment, excluding inventory for pharmacies | $I$ | CLP 15 to 40 million | CLP 25 to 50 million | Gamma |
| Capital recovery factor (10 years, 5.5%) | FRC | 0.1327 | 0.1327 | Fixed |
| Inventory, months of cost of sales | $\mu$ | 1.5 to 3.5 | 1 to 2.5 | Gamma |
| Annual cost of working capital | $r_k$ | 10% | 10% | Fixed |

*Salaries from Mi Futuro [9]; pharmacy margin informed by FNE 2019 (chains, 36% to 41%) and a business plan for an independent pharmacy (40%; fit-out and systems about CLP 14 million), drugstore share of spending by its authorised list (ACHM 2024; 47 medicines in the first municipal drugstore) and drugstore investment by ACHM 2024 (CLP 43.6 million including container and first stock); fixed costs, logistics, inventory and staff cover are the authors' assumptions; the median spending per resident (CLP 80,000) equals national spending on medicines per person. The model counts only the director (see Supplementary Note 1 for a variant with an assistant). 10,000 draws.*

#### Supplementary Note 4. Full results

Unless stated otherwise, tables use the reference scenario of Table 1 in the main text and report the mean and 95% interval of 10,000 simulations; monetary values are present values for 2027–2036 in CLP billion, discounted at 3%. Where productive recovery is not varied, the complementary NPV fixes it at 80% as a comparison anchor; the net health and access value does not depend on it.

**Supplementary Table 11. Additional events over ten years with the proposal, by replacement scenario and increase in errors without a pharmacist (half of the drug-related harm passes through pharmacies)**

| Replacement | Increase in errors without a pharmacist | ADR admissions | ADR emergency visits | ADR deaths | Poisoning admissions | Poisoning deaths | Total deaths | DALYs |
|---|---|---|---|---|---|---|---|---|
| Low | None (0%) | 0 | 0 | 0 | 0 | 0 | 0 | 0 |
| Low | Low (20%) | 392 (130 to 961) | 1,027 (328 to 2,551) | 11 (3 to 26) | 4 (3 to 7) | 0.05 (0.02 to 0.10) | 11 (3 to 26) | 194 (63 to 489) |
| Low | Reference (39%) | 765 (253 to 1,875) | 2,002 (639 to 4,974) | 21 (7 to 51) | 9 (5 to 14) | 0.09 (0.03 to 0.20) | 21 (7 to 51) | 378 (123 to 953) |
| Low | High (67%) | 1,314 (435 to 3,221) | 3,439 (1,098 to 8,546) | 36 (12 to 88) | 15 (8 to 23) | 0.16 (0.06 to 0.34) | 36 (12 to 88) | 650 (211 to 1,637) |
| Medium | None (0%) | 0 | 0 | 0 | 0 | 0 | 0 | 0 |
| Medium | Low (20%) | 1,267 (430 to 3,073) | 3,315 (1,056 to 8,271) | 35 (11 to 85) | 14 (8 to 21) | 0.15 (0.05 to 0.32) | 35 (11 to 85) | 627 (205 to 1,556) |
| Medium | Reference (39%) | 2,470 (838 to 5,993) | 6,465 (2,060 to 16,128) | 67 (22 to 166) | 28 (16 to 41) | 0.30 (0.11 to 0.63) | 68 (22 to 166) | 1,222 (400 to 3,035) |
| Medium | High (67%) | 4,244 (1,440 to 10,296) | 11,106 (3,539 to 27,707) | 116 (38 to 285) | 48 (28 to 71) | 0.52 (0.18 to 1.09) | 116 (38 to 285) | 2,099 (687 to 5,213) |
| High | None (0%) | 0 | 0 | 0 | 0 | 0 | 0 | 0 |
| High | Low (20%) | 2,550 (890 to 6,102) | 6,674 (2,210 to 16,554) | 70 (23 to 169) | 29 (18 to 42) | 0.31 (0.11 to 0.64) | 70 (24 to 170) | 1,261 (427 to 3,102) |
| High | Reference (39%) | 4,973 (1,735 to 11,898) | 13,014 (4,310 to 32,279) | 136 (46 to 330) | 56 (35 to 81) | 0.61 (0.22 to 1.26) | 136 (46 to 331) | 2,460 (833 to 6,049) |
| High | High (67%) | 8,543 (2,980 to 20,440) | 22,357 (7,404 to 55,454) | 233 (78 to 567) | 96 (60 to 139) | 1.04 (0.37 to 2.16) | 234 (79 to 568) | 4,226 (1,432 to 10,392) |

**Supplementary Table 12. Net health and access value and complementary NPV by replacement scenario and increase in errors**

| Replacement | Increase in errors | Net health and access value | Complementary NPV with 80% recovery | Simulations in which the proposal is worse (NPV) |
|---|---|---|---|---|
| Low | None (0%) | +12 (+2 to +38) | +104 (+46 to +180) | 0.0% |
| Low | Low (20%) | −12 (−49 to +19) | +80 (+15 to +157) | 0.8% |
| Low | Reference (39%) | −34 (−105 to +6) | +57 (−29 to +141) | 8.3% |
| Low | High (67%) | −68 (−187 to −8) | +24 (−103 to +120) | 27.8% |
| Medium | None (0%) | +12 (+2 to +38) | +179 (+76 to +312) | 0.0% |
| Medium | Low (20%) | −65 (−177 to −8) | +102 (−41 to +246) | 7.0% |
| Medium | Reference (39%) | −138 (−357 to −34) | +29 (−207 to +198) | 32.9% |
| Medium | High (67%) | −246 (−621 to −71) | −79 (−462 to +141) | 68.1% |
| High | None (0%) | +12 (+2 to +38) | +273 (+113 to +477) | 0.0% |
| High | Low (20%) | −143 (−366 to −37) | +117 (−144 to +353) | 14.9% |
| High | Reference (39%) | −291 (−727 to −90) | −31 (−480 to +260) | 51.1% |
| High | High (67%) | −509 (−1,255 to −164) | −248 (−1,008 to +152) | 83.4% |

**Supplementary Table 13. Complementary NPV by productive recovery, technician opportunity cost and share of harm passing through pharmacies; in brackets, simulations in which the proposal is worse (medium replacement, 39% increase in errors)**

| Share of harm through pharmacies | Productive recovery | Technician opportunity cost 50% | 75% | 100% |
|---|---|---|---|---|
| 25% | 0% | −187 (100.0%) | −248 (100.0%) | −310 (100.0%) |
| 25% | 25% | −57 (93.7%) | −119 (99.9%) | −181 (100.0%) |
| 25% | 50% | +72 (9.0%) | +10 (38.4%) | −52 (83.2%) |
| 25% | 65% | +149 (1.7%) | +88 (7.7%) | +26 (31.1%) |
| 25% | 80% | +227 (0.4%) | +165 (1.4%) | +103 (6.7%) |
| 25% | 100% | +330 (0.1%) | +268 (0.2%) | +207 (0.8%) |
| 50% | 0% | −261 (100.0%) | −323 (100.0%) | −385 (100.0%) |
| 50% | 25% | −132 (99.4%) | −194 (100.0%) | −256 (100.0%) |
| 50% | 50% | −3 (43.1%) | −65 (77.9%) | −126 (96.8%) |
| 50% | 65% | +74 (18.0%) | +13 (37.4%) | −49 (68.5%) |
| 50% | 80% | +152 (7.2%) | +90 (16.0%) | +29 (32.9%) |
| 50% | 100% | +255 (2.4%) | +194 (4.9%) | +132 (10.4%) |
| 75% | 0% | −336 (100.0%) | −398 (100.0%) | −460 (100.0%) |
| 75% | 25% | −207 (99.9%) | −269 (100.0%) | −330 (100.0%) |
| 75% | 50% | −78 (71.9%) | −140 (92.2%) | −201 (99.2%) |
| 75% | 65% | 0 (41.3%) | −62 (65.3%) | −124 (86.5%) |
| 75% | 80% | +77 (23.0%) | +15 (37.3%) | −46 (59.0%) |
| 75% | 100% | +180 (10.2%) | +119 (17.2%) | +57 (28.0%) |
| 100% | 0% | −411 (100.0%) | −473 (100.0%) | −534 (100.0%) |
| 100% | 25% | −282 (100.0%) | −344 (100.0%) | −405 (100.0%) |
| 100% | 50% | −153 (86.5%) | −214 (97.2%) | −276 (99.7%) |
| 100% | 65% | −75 (62.9%) | −137 (81.5%) | −199 (94.5%) |
| 100% | 80% | +2 (40.2%) | −59 (57.7%) | −121 (76.4%) |
| 100% | 100% | +106 (22.5%) | +44 (32.4%) | −18 (46.5%) |

**Supplementary Table 14. Increase in errors without a pharmacist beyond which the proposal has a lower complementary NPV than the current model (half of the harm passes through pharmacies; technician's opportunity cost equal to their salary)**

| Replacement | Pharmacist regains 65% of productivity | Pharmacist regains 80% | Pharmacist regains 100% |
|---|---|---|---|
| Low | 51% | 87% | 134% |
| Medium | 26% | 46% | 73% |
| High | 20% | 35% | 56% |

**Supplementary Table 15. Share of harm passing through pharmacies beyond which the proposal has a lower complementary NPV, by error scenario and value of labour (medium replacement; "Does not cross": the NPV does not change sign between 0% and 100%)**

| Productivity regained by the pharmacist | Technician's opportunity cost | Errors +20% | Errors +39% | Errors +67% |
|---|---|---|---|---|
| 65% | 50% | Does not cross | 75% | 43% |
| 65% | 75% | Does not cross | 54% | 31% |
| 65% | 100% | 66% | 34% | 19% |
| 80% | 50% | Does not cross | Does not cross | 59% |
| 80% | 75% | Does not cross | 80% | 47% |
| 80% | 100% | Does not cross | 60% | 35% |
| 100% | 50% | Does not cross | Does not cross | 79% |
| 100% | 75% | Does not cross | Does not cross | 67% |
| 100% | 100% | Does not cross | 94% | 55% |

**Supplementary Table 16. All variants evaluated on the reference scenario (complementary NPV with 80% recovery except in rows that change it)**

| Variant | Deaths over ten years | Net health and access value | Complementary NPV | Simulations in which the proposal is worse (NPV) |
|---|---|---|---|---|
| Reference | 68 | −138 (−357 to −34) | +29 (−207 to +198) | 32.9% |
| Productive recovery 0% | 68 | −138 (−357 to −34) | −385 (−629 to −226) | 100.0% |
| Productive recovery 25% | 68 | −138 (−357 to −34) | −256 (−487 to −122) | 100.0% |
| Productive recovery 50% | 68 | −138 (−357 to −34) | −126 (−353 to +5) | 96.8% |
| Productive recovery 65% | 68 | −138 (−357 to −34) | −49 (−279 to +99) | 68.5% |
| Productive recovery 100% | 68 | −138 (−357 to −34) | +132 (−118 to +338) | 10.4% |
| Technician opportunity cost 50% | 68 | −138 (−357 to −34) | +152 (−88 to +339) | 7.2% |
| No poisoning module | 67.4 | −137 (−356 to −34) | +29 (−206 to +199) | 32.6% |
| Poisonings: admissions only (no errors managed without admission, sequelae or carers' time) | 67.7 | −138 (−357 to −34) | +29 (−207 to +198) | 32.9% |
| Poisonings: strict interception criterion (3.4%) | 67.4 | −137 (−356 to −34) | +29 (−207 to +199) | 32.6% |
| Poisonings: excludes self-medication (24.8%) | 67.6 | −138 (−356 to −34) | +29 (−207 to +199) | 32.8% |
| Poisonings: case fatality not weighted by age (0.60%; 37.7 years of life lost) | 67.6 | −138 (−356 to −34) | +29 (−207 to +198) | 32.8% |
| Poisonings: adds undetermined intent | 67.8 | −138 (−357 to −34) | +28 (−207 to +198) | 33.0% |

| Variant | Deaths over ten years | Net health and access value | Complementary NPV | Simulations in which the proposal is worse (NPV) |
|---|---|---|---|---|
| Poisonings: medication-error trend (4.4% a year) | 67.8 | −138 (−357 to −34) | +28 (−207 to +198) | 33.0% |
| Poisonings: controlled medicines in intentional poisonings (+10%) | 68.2 | −139 (−358 to −35) | +27 (−209 to +197) | 33.4% |
| Poisonings: all unintentional poisonings with an assumed 8% increase (upper bound) | 70.7 | −145 (−364 to −40) | +21 (−215 to +191) | 35.6% |
| Poisonings: upper bound plus intentional poisonings | 74.5 | −157 (−376 to −50) | +10 (−227 to +180) | 40.2% |
| ADR case fatality and years of life lost not age-weighted (2.1%; 13.4) | 52 | −107 (−274 to −24) | +60 (−126 to +218) | 20.8% |
| New outlets carry the risk of the channel they replace | 67 | −137 (−355 to −34) | +29 (−206 to +199) | 32.6% |
| Includes antibiotic resistance and adverse events (exploratory) | 80 | −164 (−385 to −52) | +3 (−238 to +175) | 43.4% |
| VSL transferred from the US, elasticity 0.8 | 68 | −178 (−455 to −48) | −11 (−301 to +176) | 47.6% |
| VSL transferred from the US, elasticity 1 | 68 | −148 (−381 to −38) | +19 (−229 to +192) | 36.5% |
| Health valued as DALYs × VSLY | 68 | −55 (−152 to −3) | +112 (−22 to +254) | 4.7% |
| Weekend hour dispenses 20% of a weekday hour | 62 | −125 (−329 to −29) | +41 (−180 to +207) | 27.9% |
| Weekend hour dispenses 40% of a weekday hour | 74 | −152 (−388 to −40) | +15 (−237 to +191) | 38.1% |
| Proportional replacement across all hours | 101 | −213 (−530 to −64) | −47 (−377 to +157) | 59.6% |
| 25% of harm passes through pharmacies | 34 | −63 (−173 to −7) | +103 (−38 to +248) | 6.7% |
| 75% of harm passes through pharmacies | 101 | −213 (−539 to −60) | −46 (−385 to +157) | 59.0% |
| All harm passes through pharmacies | 135 | −288 (−723 to −85) | −121 (−561 to +120) | 76.4% |
| Case fatality 0.5% | 13 | −27 (−83 to +10) | +140 (+30 to +276) | 0.6% |
| Case fatality 1% | 25 | −52 (−142 to −3) | +115 (−13 to +255) | 3.8% |
| Case fatality 3% | 74 | −152 (−384 to −40) | +15 (−226 to +189) | 38.1% |
| Case fatality 5% | 124 | −252 (−626 to −75) | −85 (−460 to +137) | 69.8% |
| No sequelae | 68 | −135 (−350 to −33) | +31 (−200 to +200) | 31.9% |
| Adherence not valued (0 DALYs) | 68 | −139 (−358 to −36) | +27 (−208 to +197) | 33.4% |
| Adherence 0.00025 DALYs per person-year | 68 | −139 (−357 to −35) | +28 (−208 to +197) | 33.1% |
| Adherence 0.00075 DALYs per person-year | 68 | −137 (−356 to −33) | +29 (−207 to +199) | 32.6% |
| Drugstores under current rules achieve half of the new access | 68 | −144 (−362 to −43) | +23 (−211 to +191) | 35.1% |
| Discount rate 5.5% | 68 | −121 (−314 to −30) | +28 (−180 to +179) | 32.0% |
| Discount rate 0% | 68 | −163 (−416 to −41) | +30 (−246 to +226) | 34.0% |

| Variant | Deaths over ten years | Net health and access value | Complementary NPV | Simulations in which the proposal is worse (NPV) |
|---|---|---|---|---|
| Technician paid a 20% premium for directing the pharmacy | 68 | −138 (−357 to −34) | −21 (−257 to +146) | 54.4% |
| ADR baseline from 2025 discharges | 67 | −137 (−353 to −34) | +30 (−203 to +199) | 32.3% |

**Supplementary Table 17. One-way sensitivity: deaths over ten years and complementary NPV with 80% recovery at the extremes of the range evaluated (reference scenario, other parameters at their median; base: 63 deaths and CLP +39 billion)**

| Parameter | Range evaluated | Deaths | Complementary NPV |
|---|---|---|---|
| Fraction interceptable at dispensing ($f_{\text{disp}}$) | 0.05 to 0.30 | 23 to 136 | +128 to −123 |
| Baseline ADR burden ($H_0$) | 30,000 to 140,000 | 28 to 130 | +116 to −110 |
| Gross salary of the pharmacist ($W_{\text{QF}}$) | 1.8 to 2.8 million | No change | −32 to +160 |
| Case fatality of an ADR admission ($m_{\text{ADR}}$) | 0.01 to 0.05 | 23 to 116 | +119 to −68 |
| Gross salary of the technician ($W_T$) | 0.90 to 1.30 million | No change | +71 to −25 |
| Weekend dispensings per hour, as a fraction of a weekday hour ($r_{\text{ext}}$) | 0.15 to 1.00 | 55 to 93 | +56 to −29 |
| Preventable fraction ($f_{\text{prev}}$) | 0.40 to 0.65 | 48 to 78 | +71 to +4 |
| Pharmacies in operation ($N$) | 3,800 to 5,400 | No change | +11 to +71 |
| Fraction of the population without a pharmacy that gains one ($p_{\text{ent}}$) | 0 to 0.30 | 63 to 64 | +29 to +85 |
| Fraction of opening hours at the weekend ($s_{\text{ext}}$) | 0.20 to 0.35 | 71 to 52 | +20 to +63 |
| Weekly opening hours of a chain ($H_c$) | 65 to 85 | No change | +20 to +62 |
| Share of outlets belonging to chains and franchises ($P_c$) | 0.48 to 0.60 | No change | +23 to +54 |

#### Supplementary Fig. 2. Additional deaths over ten years with the proposal

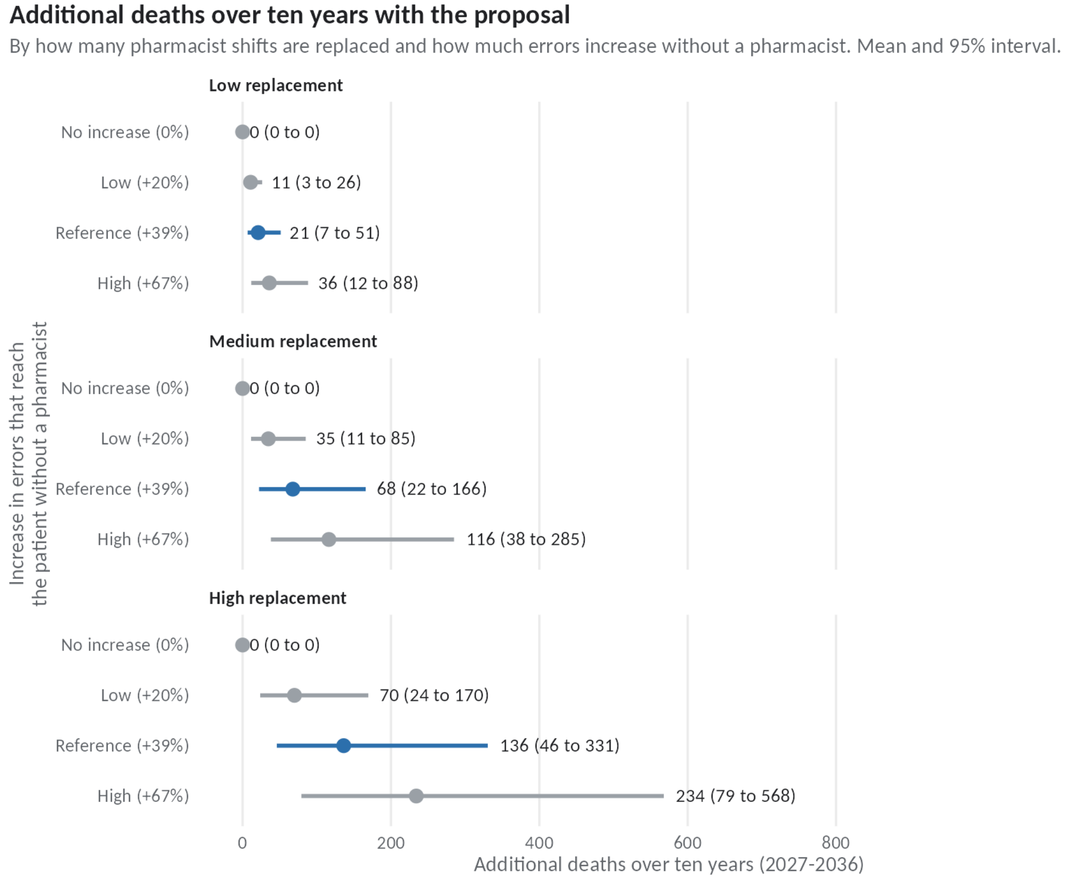


*Half of the drug-related harm passes through the affected pharmacies in every row.*

#### Supplementary Fig. 3. Complementary NPV by the share of harm passing through pharmacies, with 80% recovery

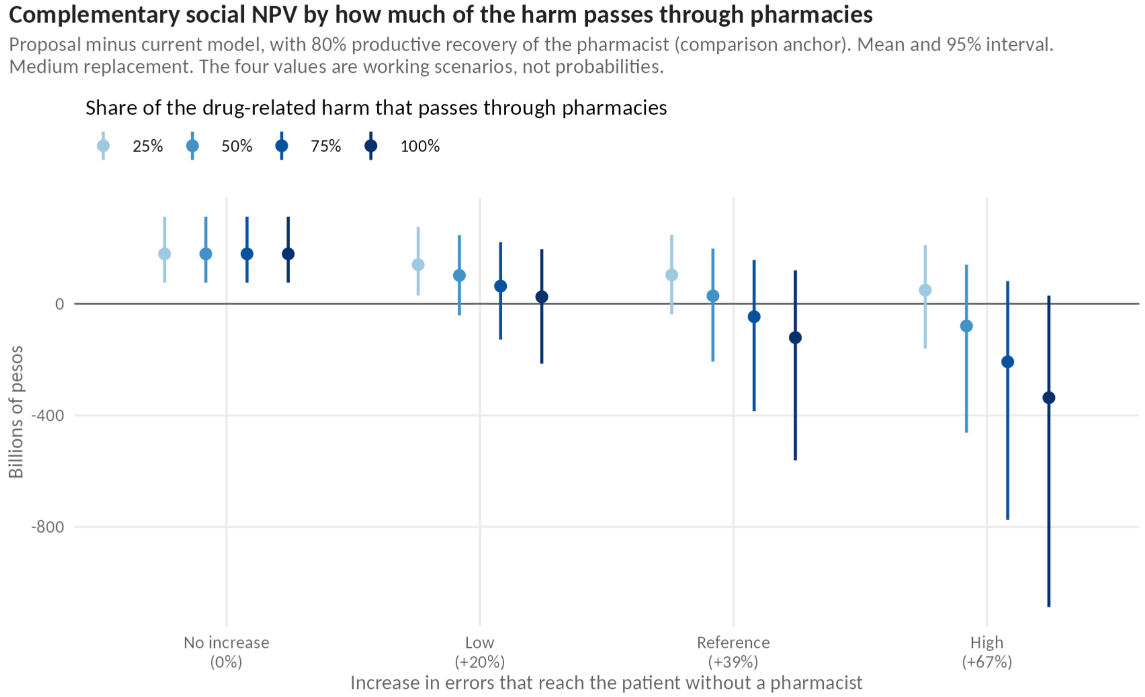


*Source: own elaboration.*

#### Supplementary Note 5. Interpreting productive recovery

Supplementary Table 18 gives the share of the present value of released labour that is still produced when re-employment is delayed (equations S14a and S14b), and the share of displaced work that must be re-employed for the expected complementary NPV of the proposal to equal that of current regulation. Delays forfeit the most heavily weighted early years and, for posts released late in the horizon, may leave no time to return before 2036. The calculation assumes a common delay; with dispersed delays the same mean delay can retain a different share of value.

**Supplementary Table 18. Value retained and share of displaced work that must be re-employed, by common delay, opportunity cost of the technician and labour cost in the new job**

| Common delay (years) | Value retained | Required share, opportunity cost 100%, same labour cost | Opportunity cost 100%, 80% of labour cost | Opportunity cost 50%, same labour cost | Opportunity cost 50%, 80% of labour cost |
|---|---|---|---|---|---|
| 0 | 100% | 74% | 93% | 51% | 63% |
| 1 | 88% | 85% | Not reached | 58% | 72% |
| 2 | 76% | 98% | Not reached | 67% | 84% |
| 3 | 64% | Not reached | Not reached | 79% | 99% |

*"Not reached": more than 100% would be needed. Reference scenario; 10,000 simulations. Maximum common delay compatible with equality: 2.1 years (opportunity cost 100%) and 4.2 years (50%).*

#### Supplementary Note 6. Selection of hospital discharges

We used the principal diagnosis and the external cause from the open hospital discharge databases for 2019 and 2023. A discharge is classified as an adverse drug event (ADE) if it has a principal diagnosis in category A1 of Stausberg and Hasford [39] (D52.1, D59.0, D59.2, D64.2, E06.4, E16.0, E23.1, E24.2, E27.3, E66.1, G21.0, G21.1, G24.0, G25.1, G25.4, G25.6, G44.4, G62.0, G72.0, H26.3, I42.7, I95.2, J70.2 to J70.4, K85.3, L10.5, L27.0, L27.1, L43.2, L56.0, L56.1, L64.0, M10.2, M32.0, M34.2, M80.4, M81.4, M83.5, M87.1, N14.0 to N14.2, R50.2, T88.6 and T88.7), haemorrhage due to anticoagulants (D68.3), toxic liver disease (K71) or an external cause in Y40 to Y59. D61, D69 and D70 are excluded, as they correspond mainly to chemotherapy given in hospital. A drug poisoning is a discharge with a principal diagnosis in T36 to T50, classified by external cause as accidental (X40 to X44), intentional (X60 to X64), of undetermined intent (Y10 to Y14) or an adverse effect in therapeutic use (Y40 to Y59); the last group is already among the selected ADEs. 2019 was used for the baseline because of data quality, not only to avoid the pandemic: it is the most recent year whose open data identify surgical discharges, and the 2023, 2024 and 2025 databases lack that variable and have more discharges of unknown age (2.5% in 2019 against 6.8% in 2024 and 6.5% in 2025). Applying the 2019 non-surgical share by age group to 2024 and 2025 discharges, distributing unknown ages proportionally and projecting to 2026 in the same way, gives a baseline 3.0% lower with 2024 and 1.1% lower with 2025 discharges; with 2025 discharges the model gives 67 additional deaths instead of 68 and a net health and access value of −137 instead of −138 billion. Case fatality is the proportion of discharges who died, with exact Clopper-Pearson intervals. For the baseline burden we used 2019 discharges without surgical intervention, excluding chapters O and P and codes Z33 to Z39, projected to 2026 by age group (0 to 19, 20 to 59, and 60 and over) with the national population projections of the National Statistics Institute (base 2024).

**Supplementary Table 19. Hospital discharges and in-hospital case fatality, 2019 and 2023**

| Group | Discharges 2019 | Case fatality 2019 (95% CI) | Discharges 2023 | Case fatality 2023 (95% CI) |
|---|---|---|---|---|
| Selected ADEs, all ages | 2,310 | 1.8% (1.3 to 2.4) | 2,773 | 2.3% (1.8 to 3.0) |
| Selected ADEs, aged 60 and over | 918 | 3.4% (2.3 to 4.8) | 1,003 | 4.1% (2.9 to 5.5) |
| Selected ADEs, under 60 | 1,392 | 0.7% (0.3 to 1.3) | 1,770 | 1.4% (0.9 to 2.0) |
| Drug poisoning (T36 to T50) | 5,559 | 0.65% (0.45 to 0.90) | 7,898 | 0.38% (0.26 to 0.54) |
| With intentional external cause (X60 to X64) | 3,315 | 0.42% (0.23 to 0.71) | 5,734 | 0.30% (0.17 to 0.47) |
| Not classified as intentional | 2,244 | 0.98% (0.62 to 1.48) | 2,164 | 0.60% (0.32 to 1.03) |
| Accidental (X40 to X44) | 704 | 0.99% (0.40 to 2.04) | 637 | 0.16% (0.00 to 0.87) |
| Undetermined intent (Y10 to Y14) | 502 | 0.20% (0.01 to 1.10) | 489 | 0.41% (0.05 to 1.47) |
| Adverse effect in therapeutic use (Y40 to Y59) | 670 | 1.49% (0.72 to 2.73) | 673 | 1.34% (0.61 to 2.52) |

Case fatality was computed by age group and weighted by the age mix of the baseline burden $H_0$, of which 65% is aged 60 or over, whereas only 39% of discharges coded as adverse drug events are (Supplementary Table 20). Years of life lost per death use the remaining life expectancy at the midpoint of each decade of age at death. The open discharge data record age in ten-year bands (under 1, 1 to 9, 10 to 19, and so on to 90 and over); every age group in the model is a union of these bands, so no interpolation was needed.

**Supplementary Table 20. Case fatality and years of life lost of admissions for adverse drug events by age group, DEIS 2019 and 2023**

| Age group | Discharges | Deaths | Case fatality | Weight in $H_0$ | Years of life lost per death |
|---|---|---|---|---|---|
| 0 to 19 | 858 | 0 | 0% | 8.9% | — |
| 20 to 59 | 2,177 | 26 | 1.19% | 26.4% | 33.8 |
| 60 and over | 1,921 | 72 | 3.75% | 64.7% | 14.6 |
| Weighted by $H_0$ | | | 2.74% (2.19 to 3.32) | 100% | 16.9 |

*127 discharges and 8 deaths of unknown age are excluded. The interval for weighted case fatality was obtained by resampling deaths in each group.*

**Supplementary Table 21. Medicine poisoning discharges by external cause, DEIS 2019 and 2023, and use in the model**

| External cause | Discharges 2019 | Discharges 2023 | Case fatality (both years) | Aged under 10 | Use in the model |
|---|---|---|---|---|---|
| Accidental (X40 to X44) | 704 | 637 | 0.60% | 63% | Baseline of the module |
| Intentional (X60 to X64) | 3,315 | 5,734 | 0.34% | 0% | Controlled-medicines variant |
| Undetermined intent (Y10 to Y14) | 502 | 489 | 0.30% | 9% | Variant |
| Adverse effect in therapeutic use (Y40 to Y59) | 670 | 673 | 1.41% | 9% | Already counted as ADRs |
| Assault (X85) | 25 | 25 | 0% | 14% | Excluded |
| Other external causes | 343 | 340 | 0.73% | 13% | Excluded |

*Other external causes are mainly unspecified accidental exposures (X59) or exposure to other substances (X49). The upper-bound variant uses all rows except the intentional one (2,204 a year) with an assumed 8% increase.*

Case fatality of accidental poisoning differs sharply by age: no one under 20 years died among 973 admissions, whereas 7 of 234 adults aged 20–59 years and 1 of 115 adults aged 60 or over died. Children make up most accidental admissions but only part of the therapeutic errors that a pharmacy could intercept, so the module weights the age-specific case fatality by the age of therapeutic errors recorded by the poison centre in 2024 [23], as is done for ADRs with the age of the baseline burden. The ages of poison-centre cases weight the DEIS figures rather than giving years of life lost directly, because the age group with most errors recorded no deaths. Since 2014, the Sanitary Code has allowed non-prescription medicines to be displayed on open shelves, with information on their use and the poison-centre telephone numbers [40]; a variant therefore excludes self-medication from the interceptable errors.

**Supplementary Table 22. Case fatality of admissions for accidental medicine poisoning by age, DEIS 2019 and 2023, and weighting by the age of therapeutic errors**

| Age group | Discharges | Deaths | Case fatality | Share of therapeutic errors, poison centre 2024 | Years of life lost per death |
|---|---|---|---|---|---|
| 0 to 9 | 846 | 0 | 0% | 45.6% | — |
| 10 to 19 | 127 | 0 | 0% | 8.7% | — |
| 20 to 59 | 234 | 7 | 2.99% | 34.4% | 42.5 |
| 60 and over | 115 | 1 | 0.87% | 11.1% | 4.0 |
| Weighted by therapeutic errors | | | 1.13% (0.44 to 2.01) | 100% | 39.2 |
| Unweighted | 1,322 | 8 | 0.60% | | |

*Discharges of unknown age are excluded. The interval was obtained by resampling deaths in each group. Poison-centre age groups (under 12, 12 to 17, 18 to 65 and over 65 years) were matched to the nearest discharge bands (0 to 9, 10 to 19, 20 to 59 and 60 and over). Because no one under 20 years died, only the boundary between 60 and 65 years affects the weighted case fatality, which the matching slightly overstates; the effect on deaths is well under one.*

**Supplementary Table 23. Therapeutic errors recorded by the national poison centre in 2024, by type, and criteria for interception at the pharmacy**

| Type of error | Cases | % | Strict criterion | Broad criterion |
|---|---|---|---|---|
| Wrong medicine | 1,140 | 34.7 | No | No |
| Wrong dose | 705 | 21.5 | No | Yes |
| Self-medication | 638 | 19.4 | No | Yes |
| Repeated administration | 435 | 13.3 | No | No |
| Wrong frequency | 98 | 3.0 | No | No |
| Wrong patient | 70 | 2.1 | No | No |
| Wrong route | 42 | 1.3 | No | No |
| Prescribing error | 41 | 1.2 | Yes | Yes |
| Dispensing error | 39 | 1.2 | Yes | Yes |
| Wrong formulation | 21 | 0.6 | Yes | Yes |
| Deteriorated medicine | 20 | 0.6 | No | No |
| Preparation or handling error | 13 | 0.4 | No | No |
| Wrong dosage form | 9 | 0.3 | Yes | Yes |
| Other (rate, duration, omission) | 12 | 0.4 | No | No |
| Total | 3,283 | 100 | 110 (3.4%) | 1,453 (44.3%) |

*Source: Silva-Silva et al. [23], table 11. The assignment of each type to a criterion is the author's, following the poison centre's proposal, and requires clinical validation. The 39 dispensing errors occurred with a pharmacist present; a poison centre does not record the errors a pharmacist intercepts.*

#### Poisonings managed without admission

The poison centre records many more therapeutic errors than are admitted: 3,283 in 2024, against about 139 admissions a year for accidental poisoning from therapeutic errors. The reference therefore applies the chain of equation S8c to the errors managed without admission, and adds sequelae and carers' time (equations S8d and S8e). Supplementary Table 24 shows the contribution of each component and its sensitivity to the assumptions. Counting these errors multiplies the number of additional poisonings by about 24 but adds little value, because most are resolved with telephone advice or a single visit and none is fatal. The centre's published reports do not give the severity of each type of error, so every error seen in a health facility is assigned two days of a moderate acute episode, and the variants halve and double that loss. These figures are a lower bound, since the centre does not receive every case.

**Supplementary Table 24. Additional poisonings from therapeutic errors over ten years, including those managed without admission (medium replacement, 39% increase in errors, half of the harm passing through the affected pharmacies)**

| Variant | Admitted | Not admitted | Seen in a facility | Under 12 | Sequelae | Carer days | DALYs | Value (CLP m) | NHA (CLP bn) |
|---|---|---|---|---|---|---|---|---|---|
| Reference | 28 | 638 (442 to 788) | 486 | 304 | 1.4 | 149 | 0.50 | 74 | −138 |
| Admissions only (no errors managed without admission, sequelae or carers' time) | 28 | 0 | 0 | 13 | 0 | 0 | 0 | 0 | −138 |
| Volume: twice the errors managed without admission (6,288 a year) | 28 | 1,276 (884 to 1,576) | 972 | 594 | 1.4 | 260 | 0.64 | 128 | −138 |
| Health loss per error managed without admission halved | 28 | 638 (442 to 788) | 486 | 304 | 1.4 | 149 | 0.43 | 71 | −138 |
| Health loss per error managed without admission doubled | 28 | 638 (442 to 788) | 486 | 304 | 1.4 | 149 | 0.64 | 80 | −138 |
| Sequelae in 10% of admitted survivors | 28 | 638 (442 to 788) | 486 | 304 | 2.8 | 149 | 0.86 | 91 | −138 |
| Carers lose 5 and 1 days | 28 | 638 (442 to 788) | 486 | 304 | 1.4 | 285 | 0.50 | 83 | −138 |
| Strict interception criterion (3.4%) | 2 | 49 (34 to 60) | 37 | 23 | 0.1 | 11 | 0.04 | 6 | −137 |
| High replacement and high increase in errors (67%) | 96 | 2,206 (1,659 to 2,589) | 1,681 | 1,050 | 4.8 | 515 | 1.73 | 256 | −509 |

*Mean (95% interval) of 10,000 simulations. Not admitted: therapeutic errors managed without admission; seen in a facility: those among them attended in a health facility; under 12: children under 12 years; DALYs and value: those added by errors managed without admission, sequelae and carers' time. NHA=net health and access value. Value is the present value, discounted at 3%, of emergency care, disability and carers' time for these events. The NHA includes them; without them it is also −138 in the reference. Children under 12 apply the share reported by the poison centre for 2024 (45.6%) to all additional poisonings.*

#### Supplementary Note 7. Pharmacy direction requirements in selected Latin American countries

Supplementary Table 25 summarises who may direct a community pharmacy, and whether a pharmacist must be present, in Chile and five other countries of the region. It describes the national rules cited and does not assess enforcement. Regulatory changes in dispensing have measurable effects in the region: enforcing prescription-only sales of antibiotics was followed by an immediate fall in antibiotic sales in Chile in 1999 [41], and by a faster decline in Mexico and a slower rise in Brazil after 2010 [42].

#### Supplementary Table 25. Who may direct a community pharmacy in selected Latin American countries

| Country | Who may direct a community pharmacy | Presence during opening hours | Source |
|---|---|---|---|
| Chile (current rules) | Pharmacist (químico farmacéutico); drugstores in municipalities without a pharmacy may be run by a technician or licensed assistant, with a restricted list | Pharmacist present throughout opening hours | Decreto Supremo 466 [43] |
| Chile (Bill 18,719-11) | Pharmacist, another health professional or a higher-level health technician trained in pharmacology and dispensing | Director present throughout opening hours; no restriction on the medicines sold | Bill 18,719-11 [44] |
| Argentina | Pharmacist as technical director, of one pharmacy only | Personal and effective attention; brief absences must be exceptional and recorded, and longer absences require a pharmacist replacement | Law 17,565, articles 25–27 [45] |
| Brazil | Pharmacist as technical lead | Pharmacist present throughout opening hours | Law 13,021/2014, article 6 [46] |
| Peru | Pharmacist (químico farmacéutico) as technical director, with assistant pharmacists | Operating without the technical director or an assistant pharmacist is an infraction | Supreme Decree 014-2011-SA, as amended by 004-2021-SA [47] |
| Colombia | Pharmacist, pharmacy technologist (regente de farmacia) or licensed drug dispenser (expendedor de drogas) for drugstores (droguerías) | Technical director required; a pharmacist is not required | Law 485 of 1998; Decree 780 of [48] |
| Mexico | A health professional (pharmacist, chemist or physician) as responsible person only where controlled medicines are sold; otherwise the owner is responsible | No professional required in pharmacies that do not sell controlled medicines | General Health Law, article 260 [49] |

#### Supplementary Note 8. CHEERS 2022 checklist

| Item | Topic | Where reported |
|---|---|---|
| 1 | Title | Title page |
| 2 | Abstract | Abstract |
| 3 | Background and objectives | Introduction |
| 4 | Health economic analysis plan | No prior plan was registered |
| 5 | Study population | Introduction; Methods (study design and comparators; access and viability) |
| 6 | Setting and location | Introduction |
| 7 | Comparators | Methods (study design and comparators) |
| 8 | Perspective | Methods (study design and comparators; separating private savings from social value); societal, with distribution by group |
| 9 | Time horizon | Methods (study design and comparators): 2027–2036 |

| Item | Topic | Where reported |
|---|---|---|
| 10 | Discount rate | Methods (study design and comparators): 3%, with 0% and 5.5% as sensitivity analyses |
| 11 | Selection of outcomes | Methods (separating private savings from social value) |
| 12 | Measurement of outcomes | Methods (dispensing without a pharmacist; adverse drug reactions; poisonings from therapeutic errors; access and viability); Supplementary Notes 1 and 6 |
| 13 | Valuation of outcomes | Methods (valuation of health; access and viability); Supplementary Note 1 |
| 14 | Measurement and valuation of resources and costs | Methods (separating private savings from social value; valuation of health); Supplementary Notes 1 and 3 |
| 15 | Currency, price date and conversion | Methods (study design and comparators: 2026 Chilean pesos); Supplementary Note 3 |
| 16 | Rationale and description of model | Methods; Supplementary Note 1 and Supplementary Fig. 1 |
| 17 | Analytics and assumptions | Methods; Table 1; Supplementary Notes 1 and 3 |
| 18 | Characterising heterogeneity | Methods (opening hours; age); Results (type of outlet) |
| 19 | Characterising distributional effects | Results (private savings and the net health and access value); Supplementary Note 1, equation S13 |
| 20 | Characterising uncertainty | Methods (uncertainty and statistical analysis); Results; Supplementary Notes 4 and 5 |
| 21 | Approach to engagement with patients and others affected by the study | None |
| 22 | Study parameters | Supplementary Note 3 |
| 23 | Summary of main results | Results; Table 2 |
| 24 | Effect of uncertainty | Results; Table 3; Supplementary Notes 4 and 5 |
| 25 | Effect of engagement with patients and others affected by the study | Not applicable |
| 26 | Study findings, limitations, generalisability and current knowledge | Discussion |
| 27 | Source of funding | Funding statement |
| 28 | Conflicts of interest | Competing interests |

.cl/fichas/130987-aportes-del-empleador-al-sistema-de-pensiones. 2025.

2016;316:2115-2125.

37. Robinson LA, Hammitt JK et al. Reference Case Guidelines for Benefit-Cost Analysis in Global Health and Development. https://content.sph.harvard.edu/wwwhsph/sites/2447/2019/05/BCA-Guidelines-May-2019.pdf. 2019.
38. Ministerio de Salud de Chile. Guía metodológica para la evaluación económica de intervenciones en salud en Chile. https://www.orasconhu.org/case/sites/default/files/files/EE_FINAL_web.pdf. 2013.
39. Stausberg J, Hasford J. Drug-related admissions and hospital-acquired adverse drug events in Germany: a longitudinal analysis from 2003 to 2007 of ICD-10-coded routine data. https://www.ncbi.nlm.nih.gov/pmc/articles/PMC3116475/. BMC Health Services Research, 2011.
40. Congreso Nacional de Chile. Ley 20.724: modifica el Código Sanitario en materia de regulación de farmacias y medicamentos. https://bcn.cl/3li15. Diario Oficial, Feb 14, 2014.
41. Bavestrello L, Cabello A, Casanova D. Impacto de medidas regulatorias en la tendencia de consumo comunitario de antibióticos en Chile. https://www.scielo.cl/scielo.php?script=sci_arttext&pid=S0034-98872002001100009. Revista Médica de Chile, 2002.
42. Santa-Ana-Tellez Y, Mantel-Teeuwisse AK, Dreser A, Leufkens HGM, Wirtz VJ. Impact of over-the-counter restrictions on antibiotic consumption in Brazil and Mexico. https://doi.org/10.1371/journal.pone.0075550. PLoS One, 2013;8:e75550.
43. Ministerio de Salud. Decreto Supremo 466, Reglamento de farmacias, droguerías, almacenes farmacéuticos, botiquines y depósitos autorizados. https://chile.justia.com/nacionales/reglamentos/decreto-n-466/gdoc; artículo 23. https://iura.cl/dto-466-minsal/23.html.
44. Boletín 18.719-11, legislative record. https://tramitacion.senado.cl/wspublico/tramitacion.php?boletin=18719. Senate of Chile.
45. Argentina. Ley 17.565: ejercicio de la actividad farmacéutica. https://www.argentina.gob.ar/normativa/nacional/norma-19424/texto. 1967 (accessed Oct 23, 2026).
46. Brazil. Lei nº 13.021, de 8 de agosto de 2014: dispõe sobre o exercício e a fiscalização das atividades farmacêuticas. https://planalto.gov.br/CCIVIL_03/_Ato2011-2014/2014/Lei/L13021.htm. 2014 (accessed Oct 23, 2026).
47. Peru, Ministerio de Salud. Decreto Supremo 004-2021-SA, que modifica el Reglamento de Establecimientos Farmacéuticos (Decreto Supremo 014-2011-SA). https://www.digemid.minsa.gob.pe/Archivos/Normatividad/2021/DS_PERUANO_004-2021-SA.pdf. 2021 (accessed Oct 23, 2026).
48. Alcaldía de Barranquilla, Secretaría de Salud. Requisitos para la inscripción y autorización para el funcionamiento de droguerías (Ley 485 de 1998; Decreto 780 de 2016). https://barranquilla.gov.co/wp-content/uploads/dlm_uploads/2025/10/requisitos-para-la-inscripcion-autorizacion-para-el-funcionamiento-de-droguerias.pdf. 2025 (accessed Oct 23, 2026).
49. Mexico. Ley General de Salud, título décimo segundo, capítulo VII, artículo 260. https://mexico.justia.com/federales/leyes/ley-general-de-salud/titulo-decimo-segundo/capitulo-vii (accessed Oct 23, 2026).
